# Blowin’ In The Wind: Long-Term Downwind Exposure to Air Pollution from Power Plants and Adult Mortality

**DOI:** 10.1101/2020.11.23.20237107

**Authors:** Shinsuke Tanaka

## Abstract

We estimate the causal effects of long-term exposure to air pollution emitted from fossil fuel power plants on adult mortality. We leverage quasi-experimental variation in daily wind patterns, which is further instrumented by the county orientation from the nearest power plant. We find that the county’s fraction of days spent downwind of plants within 20 miles in the last 10 years is associated with increased mortality from COVID-19 through the third peak in mortality in January 2021. This effect is more pronounced in fenceline communities with high poverty rates, low health insurance coverage, and low educational attainment.

---

> Yes, and how many times must a man look up Before he can see the sky?
>
> Yes, and how many ears must one man have Before he can hear people cry?
>
> Yes, and how many deaths will it take ’til he knows That too many people have died?
>
> The answer, my friend, is blowin’ in the wind The answer is blowin’ in the wind
>
> –Blowin’ in the Wind by Bob Dylan

## I. Introduction

There is a longlasting interest in the linkages between pollution from energy facilities and public health in the U.S. Fossil fuel power plants emit some of the largest amounts of hazardous pollutants into the ambient air, and concerns regarding the health impacts of exposure have resulted in a variety of public policies targeting the electric power generation industry. Existing studies suggest that people living in close proximity to fossil fuel power plants suffer a wide range of adverse health outcomes.^1^ However, the observed relationship using distance as a proxy for exposure may reflect strategic siting of power plants that confounds unobserved heterogeneities in the underlying health and/or taste-based residential sorting into the neighborhoods of plants (Heblich et al. 2016). Consequently, a recent review of the literature over the past 30 years concluded that the health costs of fossil fuel power plants remain unknown (Kravchenko and Lyerly 2018).

In this study, we conduct the first quasi-experimental investigations of the effects of long-term exposure to pollution emissions from power plants on adult mortality, while shedding light on its health consequences during a pandemic of COVID-19. To address endogeneity in the extent of pollution exposure from power plants, we leverage quasi-random variation in the exposure generated by daily wind patterns at power plants. Further, we instrument for downwind frequency using the bearing angle of the county’s orientation from the nearest power plant, whose effects are allowed to vary by geography.

We find statistically and economically significant associations between downwind frequency and the COVID-19 mortality at the county level. Our estimates suggest that the average county’s fraction of days spent downwind of power plants within 20 miles in the last 10 years led to 4.84, or 28.4%, more cumulative deaths at the first peak of the 7-day moving average deaths on April 21, 2020; 57.82, or 77.7%, at the second on August 2, 2020; and 85.52, or 44.9%, at the third on January 13, 2021, respectively, than were completely upwind counties within the same distance from plants over the same period. Further, we find that a per unit increase in the power-plant-generated long-term average PM2.5 exposure explains a 20.9% increase in cumulative COVID-19 deaths at the third peak. The robustness checks confirm that the effects are stronger in areas directly downwind than in areas lying at a greater angle from that direct line, relative to areas completely upwind within a 20-mile radius. These findings are consistent with what we find in the associations between wind patterns and PM2.5 concentrations at the monitor levels of analysis. Further analysis highlights that these effects are amplified in counties with high poverty rates, lower health insurance coverage, and low educational attainment, reinforcing the concern in light of the recent environmental justice literature that the greater burden of a pandemic is borne by people who are already at high risk (Currie 2011; Hsiang et al. 2019; Tanaka et al. Forthcoming). The falsification tests find no evidence of an association with downwind frequency of nuclear power plants or with downwind frequency for counties 20–50 miles away from power plants. In addition, we find little effect of short-term exposure to air pollution from power plants on COVID-19 mortality.

We contribute to the recent, yet a small number of, studies that exploit quasi-experimental methods for causal inference on the health burdens of fossil fuel power plants. These studies exclusively focus on infants, children, or short-term exposure. For instance, Luechinger (2014) show that the mandated installation of desulfurization systems at power plants in Germany resulted in improved SO_2_ pollution and infant mortality rates; Yang et al. (2017) and Yang and Chou (2018) find that the shutdown of a coal-fired power plant in New Jersey resulted in lower likelihood of low birthweight births; Cesura et al. (2018) show that the displacement of coal by natural gas as an energy source led to reduced mortality among adults and the elderly; and Barreca et al. (2017) document that the Acid Rain Program resulted in gradually decreasing adult mortality over time relative to reductions in air pollution in counties near regulated plants. However, there is little quasi-experimental evidence of the causal effect on adult mortality of long-term exposure to pollution emissions from power plants. We contribute new data that offer insights on spatial variation in wind patterns to establish long-term exposure to pollution emissions to certain fenceline populations living in proximity to power plants. In addition, we are not aware of any other study using quasi-experimental designs that directly estimates the mortality effects of long-term average PM2.5 concentration levels. Such estimates hold important policy implications for the cost-benefit analysis on countless regulations to curb emissions of toxic pollutants.

Our study is also one of an increasing number of studies that explain spatial variations in the effects of COVID-19 across the U.S. counties (Allcott et al. 2020; Bursztyn et al. 2020; Barrios and Hochberg 2020). The high incidence of COVID-19 mortality in low-income, minority communities has raised questions about the role air quality and pollution play in the epidemiology of COVID-19 mortality. Emerging epidemiological studies suggest that short-term exposure to air pollution exacerbates the severity of COVID-19 health outcomes (Copat et al. 2020; Ali and Islam 2020). However, evidence on whether long-term exposure to air pollution is associated with COVID-19 mortality is scarce and inconclusive due to prevailing endogeneity issues in the so-called “ecological analysis,” which links the long-term average pollution concentrations to COVID-19 mortality (Wu et al. 2020; Mendy et al. 2021; Ogen 2020). For example, Knittel and Ozaltun (2020) find that such an association suffers from an omitted variable bias, making it unclear whether the observed relationships reflect a causal effect of air pollution exposure on COVID-19 mortality or whether other factors associated with lower air quality could explain greater mortality. Further, validity of studies linking long-term pollution exposure to mortality in general (Dockery et al. 1993; Pope et al. 1995; Abbey et al. 1999; Pope et al. 2002, 2004; Miller et al. 2007; Jerrett et al. 2009; Pope et al. 2009) has been questioned with regard to the similar identification issues. We overcome such endogeneity issue by specifying pollution sources at power plants and by exploiting quasi-random wind-generated variation as well as the instrumental variable approach in measuring the long-term exposure to air pollution. Such quasi-experimental estimates are an important first step to understanding whether ongoing ambient pollution from power stations in the U.S. raises COVID-19 mortality risk in fenceline communities. Further, such data will assist U.S. policymakers to rebuild the economic recovery in a manner that effectively allocate resources to the most vulnerable regions and best prepares for future pandemics.

## II. Data

### A. Data sources

#### COVID-19 deaths

We use the publicly-available county-level cases of COVID-19 deaths from the Center for Systems Science and Engineering Coronavirus Resource Center at Johns Hopkins University (Dong et al. 2020). The data report the cumulative incidence of confirmed cases and deaths at the county-daily level in the U.S. based on various sources, including the World Health Organization (WHO), the Centers for Disease Control and Prevention (CDC), and local state health departments. We focus on the period between April 1, 2020, when the data coverage of counties is nearly complete, and January 13, 2021, when the 7-day moving average number of daily deaths was at its third peak. The number of deaths declined sharply in the following period thanks to the vaccination rollout (Online Appendix Figure A.1).

#### Power plants

We compiled the comprehensive list of all power plants that operated in 2010–2019 from various sources. First, we obtained the list of all power plants from the Emissions & Generation Resource Integration Database (eGRID). The dataset reports the geocoded addresses, as well as primary fuel sources, of almost all electric power generated in the U.S. every two years between 2010 and 2018. Where plants have switched their primary fuel sources over time, we define the primary fuel source at each plant by the fuel sources that are reported most frequently during these years. We include power plants that use coal, natural gas, and oil as the primary fuel sources. We removed from the list power plants whose sum of net generation in 2010–2019 is zero or less as reported in Energy Information Administration (EIA)-923. We further refine the plant activities by accounting for the operation status at the plant-year-month level based on the initial operation month and year and retirement month and year reported in EIA-860. In total, there are 2,740 fossil fuel power plants in the U.S. during our study period, of which 514 plants are coal-fired, and 1,642 use natural gas as their primary fuel source.

#### Meteorological data

The daily wind direction data are obtained from GRIDMET, which reports high-spatial resolution (about a 4 km by 4 km grid cell) surface meteorological data over the contiguous U.S. (Abatzoglou 2013). We match each power plant with the nearest grid cell in GRIDMET and construct the daily wind direction at each plant as measured in degrees clockwise, toward which the wind blows from power plants, normalized to be zero at northward. The average distance between power plants and the nearest grid cell is 1.0 mile, with a standard deviation of 1.1 miles. In addition, GRIDMET reports daily maximum and minimum temperatures and relative humidity, from which we construct the daily averages at the county level by taking the arithmetic averages of all grid cells that fall within each county.

#### Air quality

The daily concentrations in the ambient air of PM2.5, atmospheric particulate matters with diameters of less than 2.5 micrometers, in 2010–2019 at the monitoring stations are obtained from the publicly available Air Quality System managed by the U.S. Environmental Protection Agency (EPA). Across the U.S., more than 1,000 monitoring stations monitor PM2.5. Because these monitors do not record values every day, thus rendering inferences on the long-term average PM2.5 concentrations based on the aggregated data prone to recording frequency, we also rely on modeled PM2.5 from the study by van Donkelaar et al. (2019), which combines ground-level monitors, chemical transport models, and satellite observations to predict the monthly surface-level PM2.5 concentrations at a high-resolution of 0.01*^◦^ ×* 0.01*^◦^* for 2010–2016.^2^ We follow Wu et al. (2020) to construct the average PM2.5 levels by taking the average of all values within a county.

#### Demographics

We use the 1-year and 5-year estimates of the 2018 American Community Survey to obtain an extensive set of county characteristics. The list of individual county characteristics is presented under Table 1. We compute population density in persons per square mile using the land area from the 2010 Census.

**Table 1:** Effect of Downwind Frequency on COVID-19 Mortality

|  | 1st peak<br>Apr 21, 2020 |  | 2nd peak<br>Aug 2, 2020 |  | 3rd peak<br>Jan 13, 2021 |  |
| --- | --- | --- | --- | --- | --- | --- |
|  | OLS | 2SLS | OLS | 2SLS | OLS | 2SLS |
|  | (1) | (2) | (3) | (4) | (5) | (6) |
| Downwind | 0.778***<br>(0.297) | 1.132**<br>(0.524) | 1.482***<br>(0.357) | 1.910***<br>(0.662) | 1.149***<br>(0.307) | 1.465***<br>(0.543) |
| Controls | Y | Y | Y | Y | Y | Y |
| State FE | Y | Y | Y | Y | Y | Y |
| Mean | 17.060 | 17.060 | 74.451 | 74.451 | 190.373 | 190.373 |
| R<sup>2</sup> | 0.702 |  | 0.715 |  | 0.699 |  |

*Notes:* This table presents the effects of downwind frequency on COVID-19 mortality. The OLS model is Equation (3), and the 2SLS model is Equation (4). The independent variable is the fraction of days the county's centroid spent downwind of plants within 20 miles in the last 10 years. The sample size is 1,604 counties. The level of observations is at the county level. Controls include demographic variables (median age, gender composition, the percentage of population aged 65 and above, population, population density, the percent of whites and blacks), economic variables (the log of median household income, unemployment rate, the percent of population under the federal poverty line, median housing value), education (the percent without a high school degree, with a high school degree, and with a bachelor's degree and above), health insurance (the percentage without health insurance, with Medicare, and with Medicaid, among population aged 65 and above), geographic variables (latitude and longitudes of population-weighted centroid), the health facility capacity (the number of ICU beds), and meteorological factors (daily average precipitation, humidity, and temperature in every 5-degree Celsius bin). Additional coefficients are presented in Online Appendix Table A.3. The $F$ -statistic of excluded instruments in the first stage is 645.24. The heteroskedasticity-adjusted standard errors are reported in the parentheses.
\*\*\* $p < 0.01$ \*\* $p < 0.05$ \* $p < 0.1$

#### Hospital capacity

We collect the data on the number of intensive care unit (ICU) beds from the Homeland Infrastructure Foundation-Level Data. We aggregate data to the county level.

### B. Study population and summary statistics

The summary statistics are presented in Online Appendix Table A.1.^3^ Because the wind data are available only in the contiguous U.S., our sample in the analysis excludes Alaska and Hawaii.^4^ Our main sample is further restricted to counties whose population-weighted centroids^5^ are located within a 20-mile radius of power plants (for the reason explained below). These exclusions narrow the sample down to 1,604 counties, or 58.8% of all counties in the contiguous U.S. with the COVID-19 mortality data. In total, 190.37 people (124.02 deaths per 100,000 population) died from COVID-19 in an average county as of January 13, 2021, when the seven-day moving average of the daily deaths count was at the third peak. Overall, our sample accounts for 85.6% (305,359) of total COVID-19 deaths as of January 13, 2021 in the contiguous U.S. excluding NYC. The cumulative number of confirmed cases is 11,762.6 in an average county (18,867,128 total cases in our sample) as of January 13, 2021. Figure 1 Panel A illustrates the distributions of deaths counts in the U.S.

**Figure 1:**
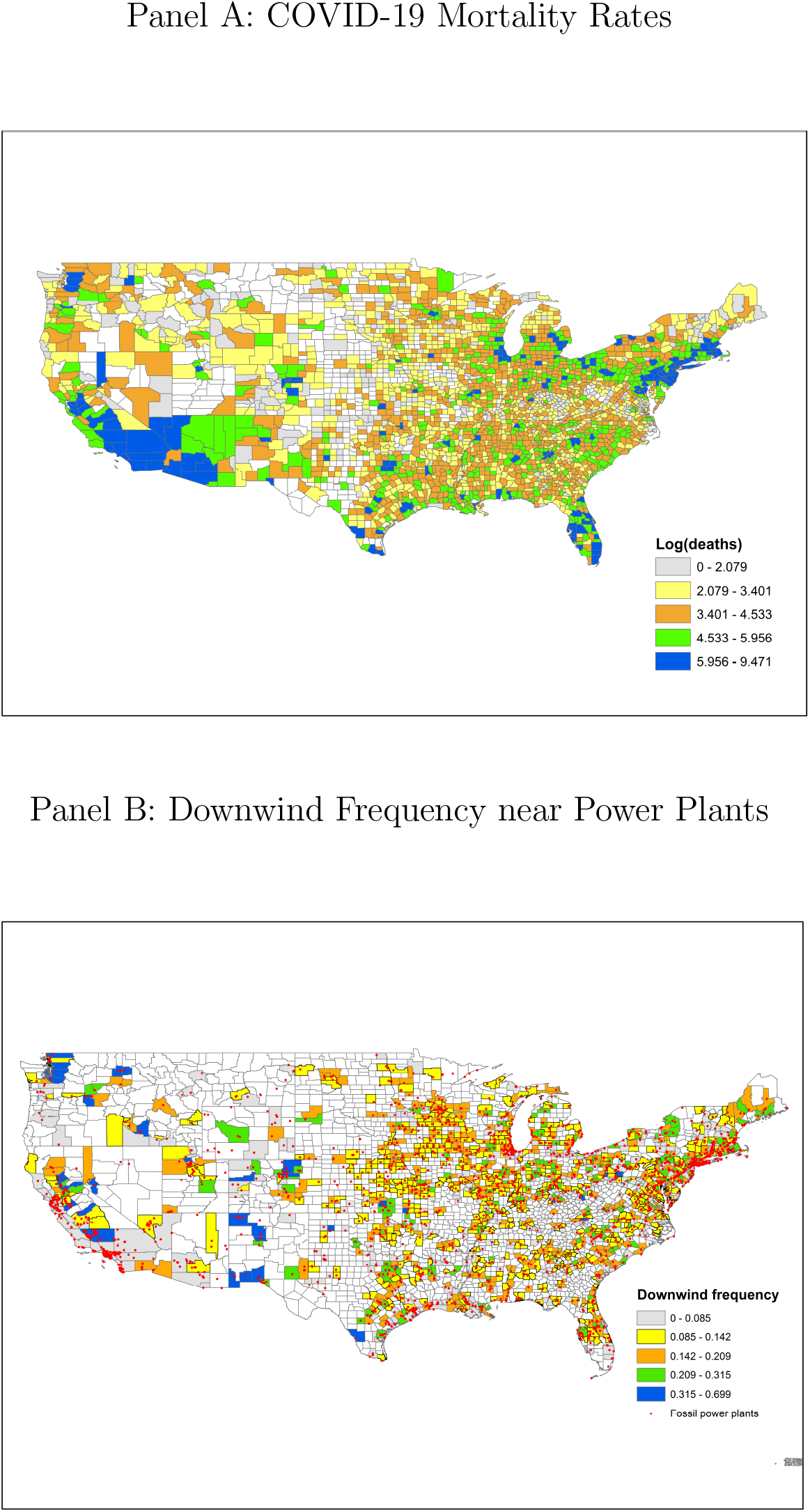
Distributions of Mortality and Downwind Frequency *Notes*: Panel A illustrates the distributions of COVID-19 deaths (log of the cumulative number of deaths plus one) as of January 13, 2021. Panel B illustrates the locations of fossil fuel power plants in the sample and downwind frequency, as measured by the fraction of days spent downwind in 2010–2019, for counties whose population-weighted centroids fall within a 20-mile radius of these power plants. Counties in white are not in the sample because mortality is not reported in Panel A and because their population-weighted centroids are over 20 miles away from power plants in Panel B.

## III. Econometric Framework

### A. The effect of downwind frequency on air quality

Our main analysis relies on the presupposition that counties downwind of power plants have greater exposure to pollutants emitted from power plants relative to counties upwind for a given distance from power plants. A number of epidemiological studies on the effects of power plants on health reviewed by Amster and Levy (2019) have found adverse health effects within a 20-mile radius. This guides us to start our main analysis below with counties within 20 miles of power plants and to empirically determine distance over which significant associations between downwind frequency and COVID-19 mortality are detected. This subsection explores whether our data support associations between downwind frequency and air quality at these hypothesized radii.

The analysis uses the daily concentrations of PM2.5 in the ambient air recorded at ground-level monitoring stations in 2010–2019. We focus on PM2.5 as a measure of air quality because these fine particulates are widely known to be most harmful to human health.^6^ The estimated model is similar to the main analysis (as described by Equation (4) below) except that the model is based on the panel data at the daily monitor level. In particular, we estimate:

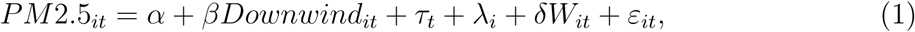

where the outcome variable is the PM2.5 concentration level in the ambient air at monitor *i* on date *t*. The independent variable of interest, *Downwind*, is a dummy variable and similarly defined as in the analysis on mortality below; the air quality monitoring station is defined as downwind of power plants on the day when it is located within 45 degrees of a ray running from the power plants to the wind direction.^7^ While the wind direction by itself is exogenous, the model includes a rich set of potential confounding variables. First, the time fixed effects, *τ_t_*, include year-month fixed effects and day-of-week fixed effects to control for seasonality effects and macroeconomic effects for given month of the year as well as for trend patterns across days of the week. Second, the inclusion of monitor fixed effects, *λ_i_*, addresses endogenous placement of monitoring stations where the EPA is concerned about high pollution by effectively comparing pollution concentrations for given monitors when they are and are not downwind. Last, the included meteorological variables, *W_it_*, are those commonly considered to affect pollution dispersion and biochemical processes of pollutant transformation, such as daily precipitation, daily average humidity, and daily average temperature in every 5-degree Celsius bin. *ε* is an idiosyncratic error. The standard errors are clustered at the monitor level.

The analysis so far uses dichotomous distinctions to define the treatment status. Instead, we now plot the marginal impacts based on the continuous measures in differences in angles between the wind direction and monitor orientation, *Angle*, distance from the nearest power plants, and the interactions of these angles and distance. In particular, we estimate:

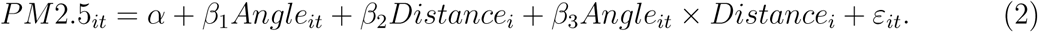

### B. The effect of downwind frequency on COVID-19 mortality

We are interested in testing whether long-term exposure to pollution emissions from power plants contributes to COVID-19 mortality. A simple comparison of counties that are near and far from power plants would generate a spurious correlation due to unobserved differences in characteristics that are correlated with distance to power plants. The ideal—though practically unfeasible—experiment to test our hypothesis would be to randomly allocate power plants across counties that are otherwise similar. Instead, our analysis makes a close approximation to such an experiment by leveraging the wind direction as the plausibly exogenous source of variation in exposure to pollution emissions from power plants across counties that are similarly close to power plants. In particular, using counties whose population-weighted centroids are within a 20-mile radius of power plants, we estimate the following specification separately for each day between April 1, 2020 and January 13, 2021:

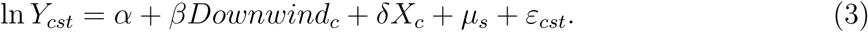

The primary outcome of interest, ln *Y_ct_*, is the log of the cumulative number of deaths plus one at county *c* on date *t*.^8^ We focus on deaths rather than confirmed cases because the latter is likely to suffer from classical and non-classical measurement errors. For instance, the number of confirmed cases crucially depends on the number of tests conducted; moreover, according to the current knowledge of COVID-19, between 17.9% and up to 50% of positive cases remain asymptomatic (Jagodnik et al. 2020; Gudbjartsson et al. 2020). In this case, a greater number of tests is likely to be conducted among high-risk populations, causing a reverse causality to bias the estimates (Borjas 2020; Schmitt-Grohé et al. 2020). On the other hand, the number of deaths is far less subject to these concerns.

The explanatory variable of interest, *Downwind_c_*, is the fraction of days the county centroid spent downwind of power plants over the 10 years between 2010–2019. Consistent with the air quality analysis above, the county is considered to be “downwind” of power plants on the day when a county’s centroid is located within 45 degrees of a ray from the power plants to the wind direction, i.e., 22.5 degrees both eastward and westward. As robustness checks, we also experiment with alternative angles to define downwind, e.g., 90 and 180 degrees. We consider all power plants within 20 miles of county centroids in constructing the dummy variable for being downwind on the daily basis between 2010 and 2019.^9^ Finally, the constructed variable is aggregated to measure the fraction of days a county centroid spent downwind of power plants in 2010–2019. The parameter of interest is *β*, which estimates the effect on the COVID-19 mortality of the fraction of days over the last 10 years spent downwind of power plants within a 20-mile radius. The identification assumption for causal inference requires that, after controlling for the covariates, a county’s fraction of days spent downwind of power plants in the neighborhood is unrelated to factors explaining the county’s mortality from COVID-19 except through air pollution.

Our richest model includes the state fixed effects, *µ_s_*, and an extensive set of county characteristics, *X_c_*. The state fixed effects help control for heterogeneities in responses to COVID-19 at the state level. For instance, states have been responsible for mitigating the effects of an outbreak through declaring states of emergency, funding and expanding COVID-19 testing, and enacting and implementing statewide stay-at-home orders and other legislation related to COVID-19. The list of individual county characteristics included is presented under Table 1. *ε* is an idiosyncratic error.

A potential threat to the identification based on the OLS framework is that low-income households may have sorted into areas with poor air quality determined by the prevailing winds over time.^10^ While we control for an extensive set of county characteristics, the analysis based on the selection on observables does not preclude a potential selection on unobservables. Thus, we employ a two stage least square (2SLS) analysis. Motivated by Deryugina et al. (2019) and Anderson (2020), we use the orientation of county centroid to the nearest fossil fuel plant as an instrument for downwind frequency and allow the effect of the bearing angle on downwind frequency to vary by geography:^11^

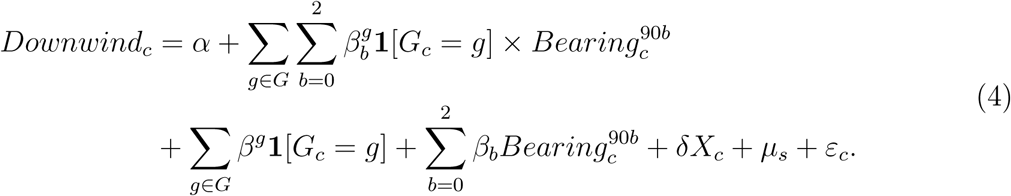

The excluded instruments are **1**[*G_c_*= *g*], *Bearing*^90*b*^_*c*_, and their interactions. The variable **1**[*G_c_* = *g*] is an indicator variable for county *c* belonging to county group *g* from the set of county groups *G*. These county groups are constructed by *k*-means cluster algorithm that partitions all U.S. counties into 90 spatial clusters based on their centroid coordinates.^12^ The variable *Bearing*^90*b*^ classifies the bearing angle of county centroid from the nearest fossil fuel power plant into four categories with a 90-degree interval of [90*b,* 90*b* + 90). The omitted category is the bearing angle in [270, 360).

Intuitively, if the winds are prevailing, the orientation of county centroids to the nearest plant affects the propensity to be downwind conditional on that these counties are spatially proximate with each other. The exclusion restriction assumes that the bearing angle of county’s orientation from the nearest power plant is not associated with adult mortality other than its influence on air quality.

Lastly, we estimate the effects of long-term average PM2.5 concentrations on COVID-19 mortality by estimating the 2SLS model. The second stage of the analysis is:

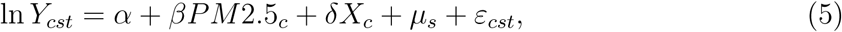

where PM2.5 is the long-term average PM2.5 concentrations. The first stage instruments the long-term average PM2.5 concentrations by the same instruments as in Equation 4:

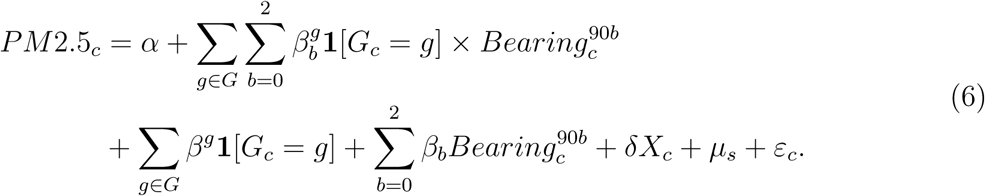

### C. The effect of short-term downwind frequency on COVID-19 mortality

The analysis thus far concerns whether the long-term exposure to pollution emissions from power plants are significantly associated with COVID-19 mortality. However, a question remains as to whether the short-term exposure has any effects on COVID-19 mortality. We answer this question based on the panel data analysis. In particular, using the same sample as in the main analysis, we employ the regression model of;

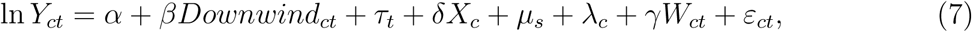

where the outcome is the log of the number of new deaths at county *c* on date *t* plus one. The main independent variable of interest, *Downwind_ct_*, is the fraction of days over the last 2 weeks the county centroid spent downwind of power plants within a 20-mile radius. We also control for the average humidity and temperature in every 5-degree Celsius bin in the past 2 weeks (*W_ct_*). The richest model includes the county fixed effects, *λ_c_*, which absorb any time invariant heterogeneities at the county level including *X_c_* and *µ_s_*. The standard errors are clustered at the county level to allow correlations in the error term at the county level over time.

The identification assumption with the county fixed effects is that after controlling for weather, day-to-day variation in wind patterns in the past 2 weeks is uncorrelated with other determinants of COVID-19 mortality.

## IV. Empirical Results

### A. Effects on air quality

We first explore whether our data support associations between downwind frequency and air quality at these hypothesized radii.^13^ The analysis uses the daily concentrations of PM2.5 in the ambient air recorded at ground-level monitoring stations in 2010–2019. We find that monitors situated closer to directly downwind of power plants are more exposed to pollutants emitted from power plants, whereas monitors situated 90–180 degrees from directly down-wind still detect pollution (Online Appendix Table A.2). For instance, we find that PM2.5 concentrations increase by 3.5% (*p* = 0.000, *n* = 1,476,800) on the days when monitors are downwind of power plants within 20 miles of power plants, whereas our estimated effect confirms no significant downwind effect (*β* = *−*0.150, *p* = 0.352, *n* = 195,616) on pollution concentrations at monitors 20 to 50 miles away.^14^ Figure 2 plots the marginal impacts based on the continuous measures in differences in angles between the wind direction and monitor orientation, distance from the nearest power plants, and the interactions of these angles and distance. The color indicates the predicted PM2.5 concentrations in ambient air with the color intensity ranging from blue (the best air quality) to red (the worst air quality), when the wind blows from north to south. The figure illustrates that there are significant spatial heterogeneities in the effects of pollution emissions from power plants depending on the monitor orientation to the wind direction, distance to a plant, and interactions of these two. It makes clear that PM2.5 concentrations are the highest in close proximity to a plant, degrade more slowly over distance in downwind than upwind, yet decay substantially at 20 miles even in downwind.

**Figure 2:**
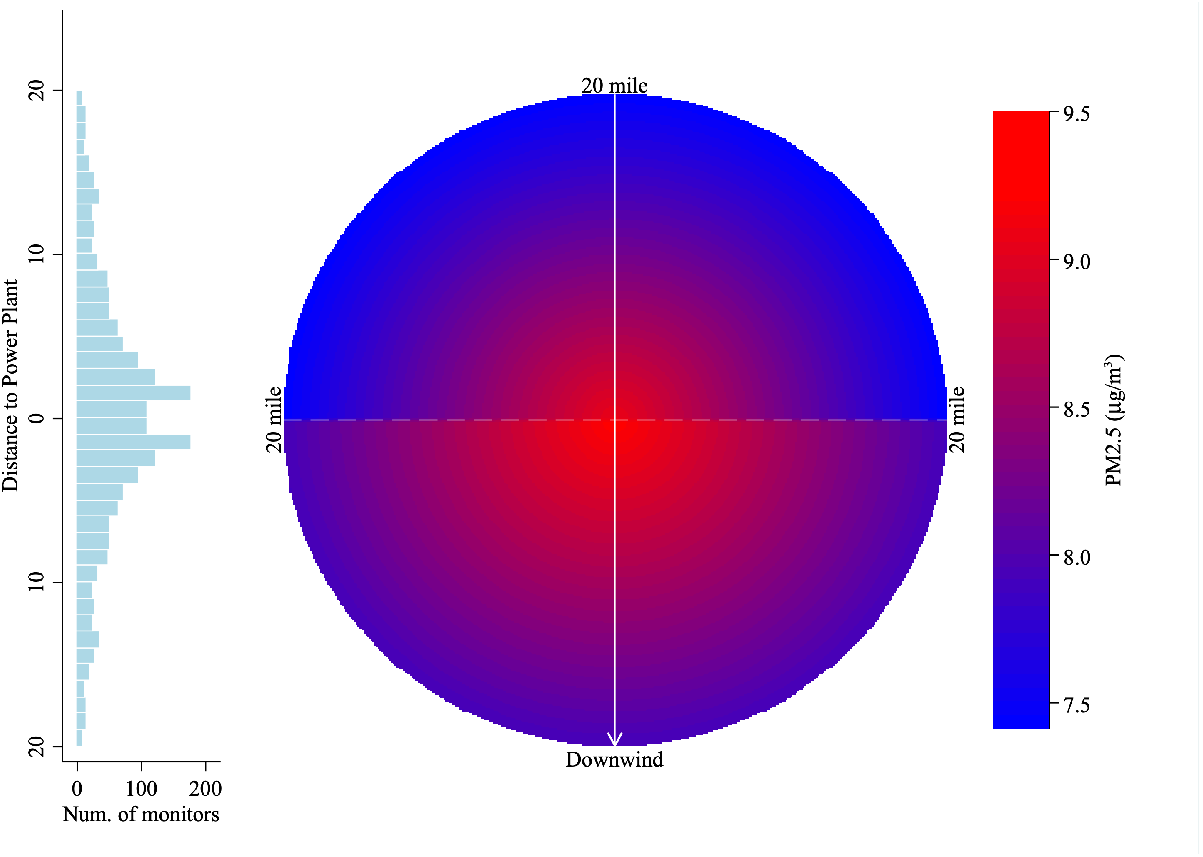
The Wind Direction and PM2.5 near Power Plants *Notes*: This figure illustrates the predicted PM2.5 concentrations in the ambient air (*µg/m*^3^) based on Equation (2) with the color intensity ranging from blue (the best air quality) to red (the worst air quality) within a 20-mile radius of a plant located at the center, when the wind blows from north to south. The left histogram indicates the number of monitors at every 1-mile distance bin from the plants.

In sum, we find stronger effects on pollution concentrations downwind than upwind up to 20 miles. Again, these pieces of evidence provide useful guidance when we analyze the mortality effects, with the caveat that they may not be directly aligned with the county-level analysis in the absence of more disaggregated mortality data. Ultimately, this leads to an empirical question that we investigate in the next subsection of whether areas at wider angles from directly downwind and farther away in distance have any associations with COVID-19 mortality.

### B. Effects on COVID-19 mortality

We now test whether long-term exposure to pollution emissions from power plants explains COVID-19 mortality. Our empirical strategy effectively compares counties with a greater fraction of days spent downwind in the last 10 years with counties with fewer such days for a given distance from power plants. Figure 1 Panel B illustrates substantial variation in downwind frequency even across spatially proximate counties. We also do not find any associations between downwind frequency and the number of fossil fuel power plants within the county, enhancing exogeneity of the wind direction with respect to the county centroid.

Figure 3 illustrates the day-by-day results using both OLS and 2SLS. Both coefficients are in the same range and suggest that downwind frequency is significantly associated with COVID-19 deaths throughout the study period. Table 1 presents the regression results on the three peak days of the 7-day moving average number of daily deaths^15^: the first peak on April 21, 2020, the second on August 2, 2020, and the third on January 13, 2021.^16^ We find statistically and economically significant associations between downwind frequency and COVID-19 mortality at the county level. The IV estimates suggest that at its first, second, and third peak, counties with the average downwind frequency of 13.5% experienced 4.84 (or 28.4%), 57.82 (77.7%), or 85.52 (44.9%), more deaths respectively than completely upwind counties within the same distance from plants.^17,18^ The estimated effect is slightly lower toward the end of the study period.

**Figure 3:**
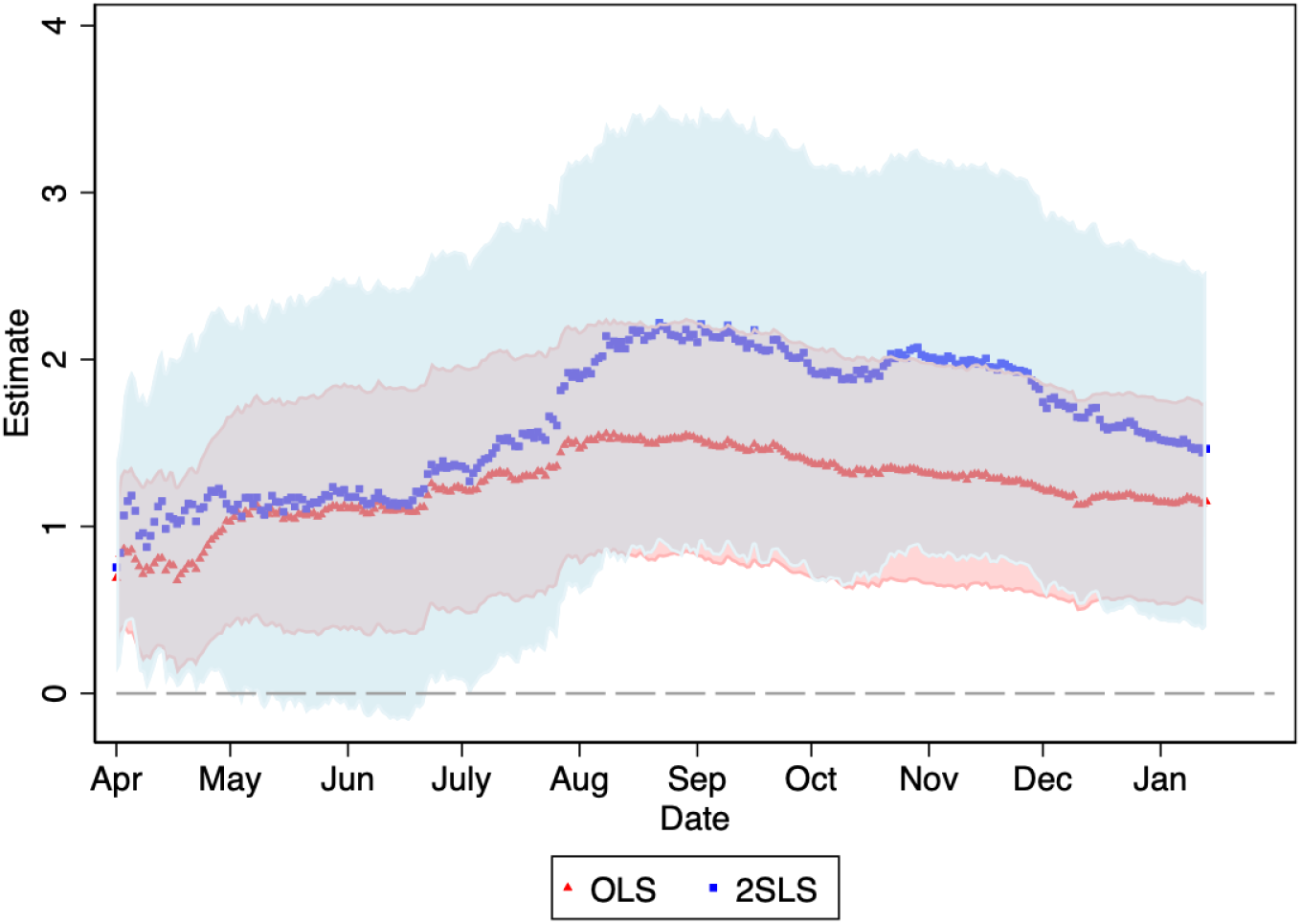
Effect of Downwind Frequency on COVID-19 Mortality *Notes*: This figure plots the coefficients and the 95% confidence interval for each day between April 1, 2020 and January 13, 2021 estimated by OLS in red and 2SLS in blue. The dependent variable is the log of the cumulative number of deaths plus one. The regressions control for the full set of county characteristics and state fixed effects. The number of observations is 1,604 for most days as illustrated by Online Appendix Figure A.4.

### C. Robustness tests

We test the robustness of the main results above to various alternative specifications.^19^ We find that the main results are robust to: i) alternative dependent variables, such as confirmed cases, the log of daily number of new deaths plus one, and mortality rates per 100,000 population; ii) alternative angles to define downwind; iii) controlling for the short-term downwind frequency to address contemporaneous effects of pollution exposure; iv) controlling for the number of power plants within 20 miles; and v) controlling for the number of industrial plants that emit toxic pollutants to address a concern that disadvantaged people often live near other hazardous sites.

Additional analyses based on Equation (7) show that the statistical correlations between downwind frequency in the past 2 weeks and COVID-19 mortality are substantially weaker and are negative when we control for the county fixed effects, suggesting that other time invariant factors related to downwind frequency in the past 2 weeks explain COVID-19 mortality.^20^ Further, downwind frequency in the past 2 weeks is not significantly associated with COVID-19 mortality and the coefficient is negative when we control for the long-term downwind frequency that remains statistically significantly associated with COVID-19 mortality. Overall, these findings are consistent with the results above that the long-term exposure to pollution emissions from power plants is an important determinant to explain across-county variation in COVID-19 mortality, whereas the short-term exposure appears to have negligible effects.

### D. Falsification tests

Next, we conduct several falsification tests.^21^ First, we find no evidence of an association between COVID-19 mortality and downwind frequency from nuclear power plants that typically do not emit air pollution hazardous to public health, suggesting that other pollution sources that are typically near power plants do not confound our estimates. Second, we find that the estimated effect is quantitatively negligible for counties 20–50 miles away from power plants, suggesting that the prevailing wind direction or county’s orientation to the power plant are not associated with COVID-19 mortality beyond what air pollution can reach within a day.

Overall, these pieces of evidence together confirm that downwind frequency is associated with the COVID-19 mortality only within a certain distance of fossil fuel power plants in which pollution exposure can be heightened by the winds.

### E. Heterogeneous effects

Next, we find that the effects are pronounced in counties with a greater share of men, greater poverty rates, a greater percent of aged population above 65 without health insurance, and a greater percent of population with less than a high school degree.^22^ These findings reinforce the concern in the large environmental justice literature that the greater burden of a pandemic is borne by people who are already at high risk (Currie 2011; Hsiang et al. 2019; Tanaka et al. Forthcoming).

### F. The effects of PM2.5 on mortality

Last, we estimate the per unit effect of long-term average PM2.5 exposure on COVID-19 mortality based on Equations (5) and (6). Table 2 provides the estimated effects on the three peak days, and Online Appendix Figure A.8 illustrates the effects on each day.^23^ Our findings are two-fold. First, we find that OLS estimates, which reproduce available evidence, e.g., Wu et al. (2020), consistently overstate the effects of PM2.5 relative to 2SLS estimates by nearly 50%. Since existing studies on the relationship between long-term exposure to air pollution and mortality exclusively rely on such so-called ecological analysis based on a similar selection-on-observables design, our finding sheds light on new evidence based on a quasi-experimental design in identifying causality. Second, while the effects of per unit PM2.5 are lower and negligible at the beginning, the estimates quickly become large and converge to a statistically significant estimate of around 0.180. At the third peak, we find that a unit increase in long-term average PM2.5 leads to 20.9% or 39.8 more COVID-19 deaths.

**Table 2:** Effect of Long-term Average PM2.5 on COVID-19 Mortality

|  | 1st peak<br>Apr 21, 2020 |  | 2nd peak<br>Aug 2, 2020 |  | 3rd peak<br>Jan 13, 2021 |  |
| --- | --- | --- | --- | --- | --- | --- |
|  | OLS | 2SLS | OLS | 2SLS | OLS | 2SLS |
|  | (1) | (2) | (3) | (4) | (5) | (6) |
| PM2.5 | 0.136***<br>(0.027) | 0.037<br>(0.036) | 0.271***<br>(0.033) | 0.180***<br>(0.045) | 0.254***<br>(0.026) | 0.190***<br>(0.035) |
| Controls | Y | Y | Y | Y | Y | Y |
| State FE | Y | Y | Y | Y | Y | Y |
| Mean | 17.060 | 17.060 | 74.451 | 74.451 | 190.373 | 190.373 |
| R<sup>2</sup> | 0.707 |  | 0.726 |  | 0.717 |  |

*Notes:* This table presents the effects of the long-term average PM2.5 on COVID-19 mortality. The OLS model is Equation (5), and the 2SLS model is Equation (6). The independent variable is the average PM2.5 concentrations in the ambient air between 2010 and 2016. The sample include 1,596 counties whose population-weighted centroids are located within 20 miles of fossil fuel power plants. The level of observations is at the county level. The same controls are included as in Table 1. Additional coefficients on the daily basis through the study period are presented in Online Appendix Figure A.8. The $F$ -statistic of excluded instruments in the first stage is 94.0. The heteroskedasticity-adjusted standard errors are reported in the parentheses.
\*\*\* $p < 0.01$ \*\* $p < 0.05$ \* $p < 0.1$

## V. Conclusions

This study presents the first evidence that substantiates the health costs of long-term exposure to pollution emissions from fossil fuel power plants in the context of COVID-19. These findings hold important policy implications for rebuilding the U.S. energy system in the aftermath of the pandemic. In particular, they suggest the possibility that investments in clean energy could contribute to building a society more resilient to airborne disease and thereby less impacted by pandemics. These results raise important questions about future regulations of older fossil fuel power plants in the U.S. and what must be studied in determining their costs to society and environmental impacts.

Further, our findings highlight that the substantial health burdens of pollution from power plants are disproportionately borne by certain fenceline communities. These findings call attention to the public health risks that unfettered pollution from energy facilities poses to fenceline communities. This kind of epidemiological study is particularly pressing in light of considerations of environmental justice and social equity in the U.S. To quote a recent statement by the United Nations Sectary-General, “[F]iscal firepower must shift economies from grey to green, making societies and people more resilient through a transition that is fair to all and leaves no one behind”.^24^

Further study could be helpful in determining what facilities should be prioritized for closure and replaced with cleaner energy sources as part of energy policy and future economic stimulus spending to reduce the impacts of airborne illness in the U.S. particularly among the most vulnerable regions as identified in this study.

## Data Availability

All data used in this analysis are free and publicly available and will be available upon publication from the corresponding author.

## Online Appendix

### Overview

This Online Appendix provides supplementary materials to what is presented in the main text. Section A reviews the extensive literature on the health burden of power plants. Section B presents the detailed summary statistics of the sample used in the main analysis. Section C presents additional results on the effects of downwind frequency on air quality. Section D.1 presents additional results on the effects of downwind frequency on COVID-19 mortality, Section D.2 presents evidence from the series of the robustness checks, Section D.3 presents results from the falsification tests, Section D.4 presents the heterogeneities in the effects, and Section D.5 presents the effect of short-term exposure to air pollution from power plants on COVID-19 mortality, and finally, Section D.6 presents additional evidence on the per unit effects of long-term PM2.5 exposure on COVID-19 mortality.

#### A. The Health Burden of Power Plants

The electricity generation industry remains one of the largest pollution emission sources, as combusting fossil fuels, such as coal, natural gas, and petroleum, continues to be the largest energy source for electricity generation. In the U.S., fossil fuels accounted for 61% of electricity generation in 2019 (U.S. Energy Information Administration 2020). Hazardous pollutants released from fossil fuel power plants include particulate matters (PM), sulfur dioxide (SO_2_), nitrogen oxides (NO_X_), lead, mercury and other toxic metals, carbon monoxide, volatile organic compounds, and arsenic. Once emitted, various meteorological factors, such as wind direction and velocity, precipitation, humidity, and temperature, determine the processes of dispersion, transportation, transformation, degradation, and decay.

There is ample convincing evidence that exposure to these pollutants imposes substantial health risks on the worldwide population. The World Health Organization (WHO) estimates that about seven million people die every year across the globe from pollution in both outdoor and indoor air. Both short-term and long-term exposure to fine particles in the air can cause elevated health risks, such as respiratory infections, ischemic heart diseases, strokes, lung cancer, and infant mortality. For example, a large body of literature substantiates the effects of short-term exposure to air pollution on fetal and infant health (Chay and Greenstone 2003a; Chay and Greenstone 2003b; Currie and Neidell 2005; Currie et al. 2009; Jayachandran 2009; Currie and Walker 2011; Tanaka 2015; Knittel et al. 2016; Arceo-Gomez et al. 2016;Currie et al. 2015); of contemporaneous exposure on child and adult health (Moretti and Neidell 2011; Schlenker and Walker 2015; Ward 2015; Gupta and Spears 2017; Deryugina et al. 2019); and of long-term exposure on adult health and mortality (Dockery et al. 1993; Pope et al. 2002; Chen et al. 2013; Barreca et al. 2017; Anderson 2020).

Not surprisingly, the electric power industry has been subject to countless regulations to curb emissions of these toxic pollutants.^1^ These regulatory actions reflect widespread consensus that people living in close proximity to fossil fuel power plants suffer a wide range of adverse health outcomes (Schneider and Banks 2010; Liu et al. 2012; Ha et al. 2015; Amster and Levy 2019). However, the observed relationship using distance as a proxy for exposure may reflect strategic siting of power plants that confounds unobserved heterogeneities in the underlying health or taste-based residential sorting into the neighborhoods of plants (Heblich et al. 2016). A smaller number of studies exploit quasi-experimental methods to identify the health burdens of exposure to air pollution from power plants. For instance, Luechinger (2014) show that the mandated installation of desulfurization systems at power plants in Germany resulted in improved SO_2_ pollution and infant mortality rates; Yang et al. (2017) and Yang and Chou (2018) find that the shutdown of a coal-fired power plant in New Jersey resulted in lower likelihood of low birthweight births; Cesura et al. (2018) show that the displacement of coal by natural gas as energy source led to reduced mortality among adults and the elderly; and Barreca et al. (2017) document that the Acid Rain Program resulted in gradually decreasing adult mortality over time relative to reductions in air pollution in counties near regulated plants. However, there still exists little quasi-experimental evidence of the causal effect on adult mortality of long-term exposure to pollution emissions from power plants. We contribute new data that offer insights on spatial variation in wind patterns to establish long-term exposure to pollution emissions to certain fence line populations living in proximity to power plants. In addition, we are not aware of any other study using quasi-experimental designs that directly estimates the mortality effects of long-term average PM2.5 concentration levels. Such estimates hold important policy implications for the cost-benefit analysis on countless regulations to curb emissions of toxic pollutants.

#### B. Data

Table A.1 presents the summary statistics for key variables in the analysis. Because the wind data are available only in the contiguous U.S., our sample in the analysis excludes Alaska and Hawaii. We also drop New York City as there is an obvious error in the number; specifically, the cumulative number of confirmed cases (deaths) decreases from 233,969 (23,689) on August 30, 2020 to 30,874 (3,170) on August 31, 2020, and starts increasing from there onward, reaching cumulative counts of only 85,779 (3,586) even on January 31, 2021. Our main sample is further restricted to counties whose population-weighted centroids are located within a 20-mile radius of power plants (for the reason explained below). These exclusions narrow the sample down to 1,604 counties, or 58.8% of all counties in the contiguous U.S. with the COVID-19 mortality data. Panel A presents the COVID-19 data as of January 13, 2021, when the seven-day moving average of the daily deaths count was at the third peak. The average number of deaths in our sample is 190.37 per county on January 13, 2021. This is equivalent to 124.02 deaths per 100,000 people. Overall, our sample accounts for 85.6% (305,359) of total COVID-19 deaths as of January 13, 2021 in the contiguous U.S. excluding NYC. The cumulative number of confirmed cases is 11,762.6 in an average county (18,867,128 total cases in our sample) as of January 13, 2021. Figure 1 Panel A illustrates the distributions of deaths counts in the U.S.

Panel B presents information related to fossil fuel power plants. The average fraction of days for counties spent downwind within 45 degrees is 13.5% with a standard deviation of 0.074, whereas the corresponding figures within 90 degrees and 180 degrees increase to 25.6% and 49.6%, respectively.^2^ The average distance to the nearest power plant from the county’s population-weighted centroid is about 9.30 miles, and on average there are 3.0 fossil fuel power plants within a 20-miles radius of county’s population-weighted centroid. Figure 1 Panel B in the main text illustrates the locations of fossil fuel power plants and downwind frequency for counties whose population-weighted centroids fall within a 20-mile radius of power plants. Clearly, the wind direction varies substantially even across spatially proximate counties. We also do not find any associations between downwind frequency and the number of fossil fuel power plants within the county, enhancing exogeneity of the wind direction with respect to the county centroid.

Panel C presents county characteristics. The average population of counties is around 170,000. About 17.1% of the population are aged 65 and above, those most vulnerable to COVID-19. The sample locations are reasonably uniformly distributed across the U.S. (Figure 1 Panel A in the main text).

Panel D presents information on air quality at the level of daily monitoring stations. Notably, the average concentrations of PM2.5 in the ambient air is 8.730 microgram per cubic meter (*µg/m*^3^), which is substantially lower than the 24-hour standard of 35 *µg/m*^3^ mandated by the National Ambient Air Quality Standards. Although these standards set the maximum allowable limits on pollution concentrations to protect public health, studies have found that pollution concentrations even below these EPA mandates are harmful to human health (Schlenker and Walker 2015; Currie et al. 2015).

#### C. Effects of Downwind Frequency on Air Quality

We first explore whether our data support associations between downwind frequency and air quality at these hypothesized radii. The analysis uses the daily concentrations of PM2.5 in the ambient air recorded at ground-level monitoring stations in 2010–2019. We focus on PM2.5 as a measure of air quality because these fine particulates are widely known to be most harmful to human health.

Table A.2 presents the results based on Equation (1) in the main text. Column (1) reports the bivariate association between PM2.5 concentrations and being downwind. The estimated coefficient implies that PM2.5 concentrations are higher by 0.307 *µg/m*^3^ on the days when monitors are downwind of power plants. This corresponds to an approximate 3.5% increase from the mean value.

Column (2) includes the time fixed effects, and Column (3) adds weather variables. In all models, the estimated effects remain statistically significant at the 1% level. Column (4) compares downwind monitors at various angles with those beyond these angles, e.g., upwind monitors. As expected, monitors situated closer to directly downwind of power plants are more exposed to pollutants emitted from power plants, whereas monitors situated 90–180 degrees from directly downwind still detect pollution.

Column (5) considers monitors situated 20–50 miles away from power plants. Note that the number of monitors is substantially lower in these areas although they cover more than 5 times the land area. Since the EPA typically places monitors near pollution sources, e.g. power plants, the number of monitors by itself reflects recognition that the population within 20 miles of power plants is at substantially greater risk. Our estimated effect confirms no significant downwind effect on pollution concentrations at monitors 20 to 50 miles away.

Note that the analysis thus far exploits variation in the downwind frequency at the daily level. Pollutants can travel over days, and areas farther downwind may or may not experience a lag in exposure to pollution emissions depending on the wind direction in the following days. Ultimately, it is an empirical question whether people farther down the wind are also exposed to pollution. Below, we still investigate the effect on mortality beyond 20 miles to infer whether pollution may potentially travel farther.

The analysis so far uses dichotomous distinctions to define the treatment status. Instead, we now plot the marginal impacts based on the continuous measures in differences in angles between the wind direction and monitor orientation, distance from the nearest power plants, and the interactions of these angles and distance.^3^

Figure 2 in the main text plots adjusted predictions within a 20-mile radius of a plant located at the center based on Equation (2) in the main text. The color indicates the predicted PM2.5 concentrations in ambient air with the color intensity ranging from blue (the best air quality) to red (the worst air quality), when the wind blows from north to south. The figure illustrates that there are significant spatial heterogeneities in the effects of pollution emissions from power plants depending on the monitor orientation to the wind direction, distance to a plant, and interactions of these two. The figure makes clear that PM2.5 concentrations are the highest in close proximity to a plant, degrade more slowly over distance in downwind than upwind, yet decay substantially at 20 miles even in downwind. These findings reinforce the regression results.

In sum, we find stronger effects on pollution concentrations downwind than upwind up to 20 miles. Again, these pieces of evidence provide useful guidance when we analyze the mortality effects, with the caveat that they may not be directly aligned with the county-level analysis in the absence of more disaggregated mortality data. Ultimately, this leads to an empirical question that we investigate in the next section of whether areas at wider angles from directly downwind and farther away in distance have any associations with COVID-19 mortality.

#### D. Effects on COVID-19 Mortality

##### 1. Main results

The primary goal of this paper is to characterize the associations between downwind frequency with respect to power plants and COVID-19 mortality. Table A.3 presents the regression results using the specification outlined in Equations (3) and (4) in the main text, including all coefficients besides what is already presented in Table 1 in the main text. The analysis controls for the set of county characteristics as well as state fixed effects. We report the estimated effects using both OLS and 2SLS on the three peak days: the first peak on April 21, 2020, the second on August 2, 2020, and the third on January 13, 2021. We find statistically and economically significant associations between downwind frequency and COVID-19 mortality at the county level. At its first, second, and third peak, counties with average downwind frequency (0.135) experienced 2.71 (or 15.9%), 57.82 (77.7%), 85.52 (44.9%), more deaths respectively than completely upwind counties within the same distance from plants. An alternative way to interpret these estimates is that each additional 10 days spent downwind in the last ten years led to 0.10, 1.17, or 1.73 more deaths at the first, second, and third peak, respectively. The estimated effect is found to be slightly lower toward the end of the study period.

##### 2. Robustness

We test the robustness of the main results above to various alternative specifications. The results are presented in Table A.4 Panel A. First, we consider alternative dependent variables. Specifically, the first row considers the log of the number of cases plus one as the outcome, the second the log of new daily deaths plus one, and the third the mortality rates per 100,000 population.^4^ The 2SLS estimates suggest that counties with average downwind frequency (0.135) experienced 185.4 (48.7%), 2,398.4 (100.8%), and 3943.5 (33.5%) more cases, 0.177 (16.6%), 0.030 (12.4%), 0.578 (25.3%) more daily new deaths, and 4.66 (88.7%), 9.06 (31.0%), 2.840 (2.29%) more deaths per 100,000 population at the first, second, and third peak days respectively than completely upwind counties within the same distance from plants.^5^

Next, we consider alternative angles to define downwind. In particular we consider 90 degrees, or 45 degrees on both the eastward and westward sides of a ray from power plants to wind direction, and 180 degrees, or the south semicircle when the wind blows south, to construct downwind frequency. We find that the estimated effects are slightly lower within 90-degree angles, and the effects are even lower within 180-degree angles, suggesting that the effects consistently decrease as differences in angle between the wind direction and county centroid increase.

Next, we control for the short-term downwind frequency to disentangle the effect of short-term exposure from those of long-term exposure. In particular, we include downwind frequency during the last 14 days for each day. The estimated effect of the long-term downwind frequency remains similar. In Section 5 below, we further examine the effect of short-term exposure on COVID-19 mortality based on the panel data. The finding consistently suggests that the long-term exposure has significant impacts on COVID-19 mortality, whereas the effect of the short-term exposure is negligible.

Next, we control for the number of power plants within 20 miles. This variable is not included in the main analysis because downwind frequency is likely a function not only of wind direction but also of the number of power plants in the neighborhood (although our data do not show any associations between downwind frequency and the number of power plants owing to exogeneity in the wind direction with respect to the county centroids). Yet the inclusion of this variable helps illustrate an effect of downwind exposure independent of the number of power plants. The estimated effect is robust to such inclusion.

Last, we control for the number of industrial plants that emit toxic pollutants to control for other pollution emission sources. These data are obtained from the Toxic Release Inventory (TRI), which reports the locations of these plants as well as their types and volumes of emissions. Although plants that meet certain requirements are mandated to report their emissions, the amounts are self-reported without formal verification processes, making these values subject to substantial errors (de Marchi and Ham 2006; Koehler and Spengler 2007). We follow Currie et al. (2015) by including all facilities that have emitted any toxic pollutants in the study period from 2010 up to 2018, the most recent year when the data are available and counting the number of such facilities at the county level. The estimated effect of downwind frequency is unchanged.

##### 3. Falsification tests

In this section, we present results from the set of falsification tests. First, we test the effects on COVID-19 mortality of downwind frequency from nuclear power plants.^6^ Unlike fossil fuel power plants, nuclear power plants do not produce air pollution hazardous to public health. Figure A.7 shows that none of the estimates is statistically significantly different from zero throughout the study period, and most estimates are negative. Table A.4 Panel B presents the consistent results at the three peak days.^7^ The evidence suggests that environmental disamenities that are typically associated with power plants and downwind frequency do not explain the main results.

Second, we consider counties 20–50 miles away from power plants. Although there is likely to be a wide variation, the literature has typically considered those within a 20-mile radius of power plants at risk of high pollution exposure. Further, our analysis of air pollution also finds that monitors detected greater pollution concentrations on a day spent downwind only within 20 miles. Nonetheless, whether counties 20–50 miles away from power plants are exposed to pollution emitted from power plants remains an open for two reasons.

First, the analysis in the main text exploits variation in the downwind frequency at the daily level. Pollutants can travel over days, and areas farther downwind may or may not experience a lag in exposure to pollution emissions depending on the wind direction in the following days, as discussed above. Second, unlike stationary pollution monitors, there is a large measurement error in areas of exposure for people because i) we do not know exact residential locations for each individual, and ii) people move around. This may lead counties 20–50 miles away from power plants to be exposed to pollution from power plants. Ultimately, it is an empirical question of whether people farther down the wind are also exposed to pollution.

We find that the estimated effect is quantitatively negligible for counties 20–50 miles away from power plants, suggesting that the associations between the downwind frequency and COVID-19 mortality decay substantially after 20 miles (Figure A.7 for each day and Table A.4 Panel B for the three peak days).

Overall, these pieces of evidence together confirm that greater downwind frequency is by no means correlated with heterogeneities other than pollution exposure that would explain greater COVID-19 mortality. Rather, downwind frequency is associated with the COVID-19 mortality only within a certain distance of fossil fuel power plants in which pollution exposure can be heightened by the winds.

##### 4. Heterogeneities in the effects

Table A.5 explores heterogeneities in the effects of downwind frequency on COVID-19 mortality across various subsample of counties determined by being above or below the median of several county characteristics.^8^ The analysis includes the set of county characteristics and state fixed effects. We find that the effects are pronounced in counties with a greater share of men, the greater poverty rate, a greater percent of aged population above 65 without health insurance, and a greater percent of population with less than a high school degree. On the other hand, the heterogeneity with respect of percent of Black population is less clear. These findings highlight that the greater burden of a pandemic is borne by already high-risk subjects.

##### 5. The effect of short-term downwind frequency on COVID-19 mortality

The results based on Equation (7) in the main text are presented in Table A.6. Column (1) shows that downwind frequency in the past 2 weeks is significantly associated with COVID-19 mortality. Column (2) adds distance to a plant as a control. The estimated effect decreases by about 40%. Column (3) includes the same set of controls that are included in the main analysis, and Column (4) additionally controls for the state fixed effects. The estimated effects become even smaller yet remain statistically significant. Column (5) includes the county fixed effects, which control for any time invariant heterogeneities that explain differences in COVID-19 mortality across counties. The point estimate becomes negative and statistically significant. This suggests that other time invariant factors related to downwind frequency in the past 2 weeks explain COVID-19 mortality. Column (6) additionally includes downwind frequency in the last 10 years to Column (4), and we find that the long-term downwind frequency has significant effects on daily COVID-19 mortality, whereas the point estimate of downwind frequency in the last 2 weeks remains negative and statistically indistinguishable from zero.

Overall, these findings suggest that the long-term exposure to pollution emissions from power plants is an important determinant to explain across-county variation in COVID-19 mortality, whereas the short-term exposure appears to have negligible effects.

##### 6. The per unit effect of long-term average PM2.5 on mortality

Figure A.8 illustrate the effects of long-term average PM2.5 concentrations on COVID-19 mortality on each day throughout our study period. The OLS model is based on Equation 5, and the 2SLS model is based on Equation 6 in the main text.

**Figure A.1:**
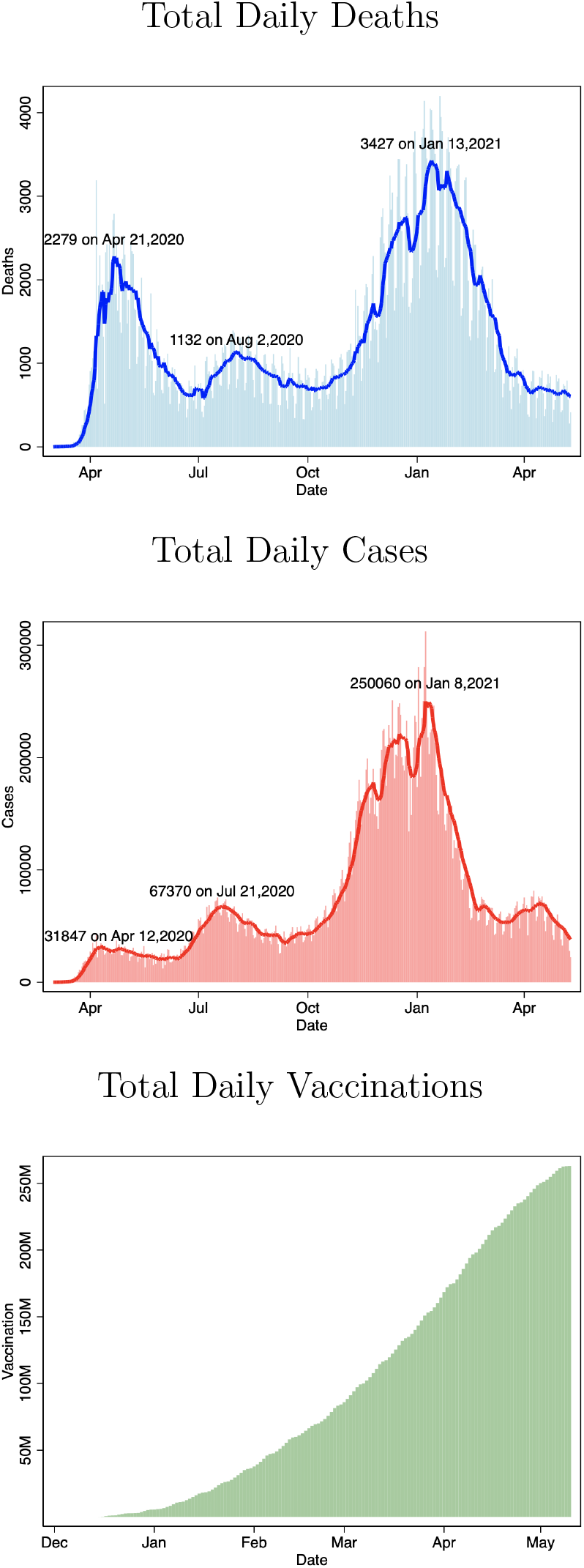
COVID-19 Trends in the U.S. *Notes*: The top two figures show the number of daily deaths and cases across the U.S., respectively. The solid lines indicate the 7-day moving averages. I noted the three peak days and the associated numbers. The bottom figure shows the number of daily doses administered across the U.S. starting December 2020. *Date source*: CDC COVID Data Tracker.

**Figure A.2:**
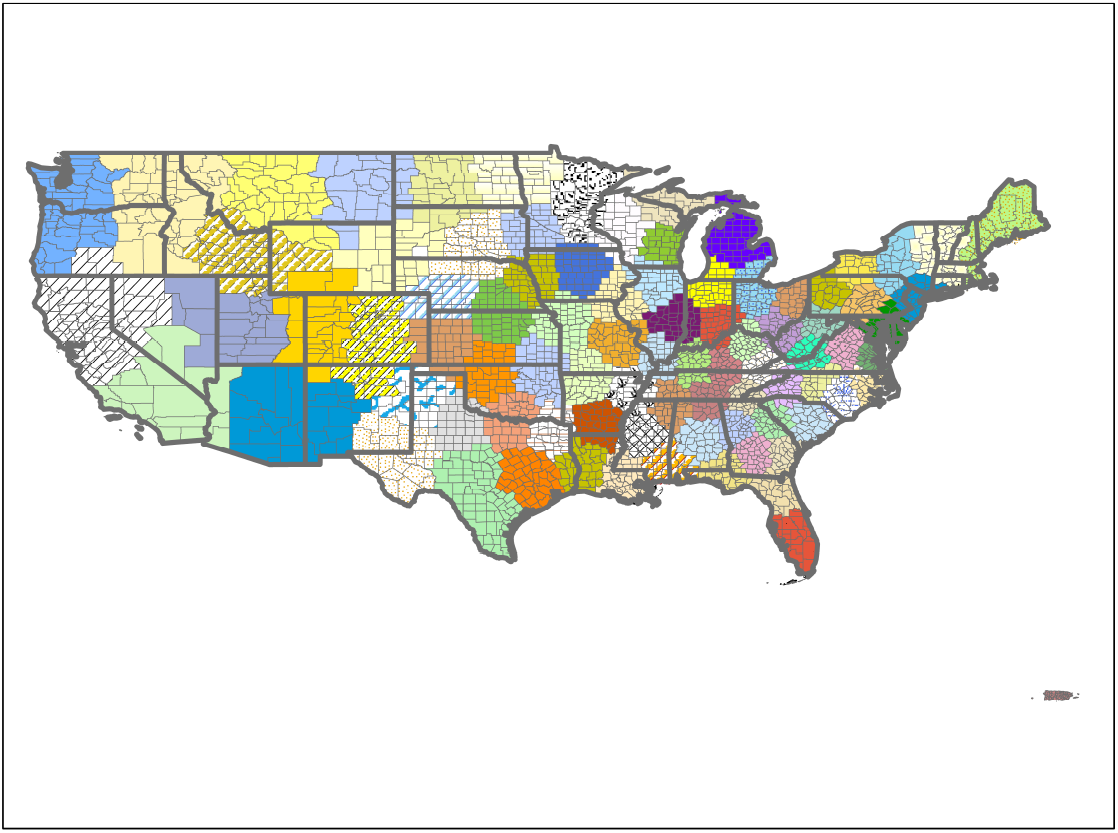
The Distribution of County Clusters *Notes*: This figure illustrates the distribution of 90 county clusters constructed by *k*-means cluster algorithm using county centroids. Each color represents a group, but because making 90 different color schemes is practically not meaningful, groups with a similar color should be considered as separate groups unless they are spatially adjacent with each other. The thick lines indicate state borders, and the thin lines indicate county borders.

**Figure A.3:**
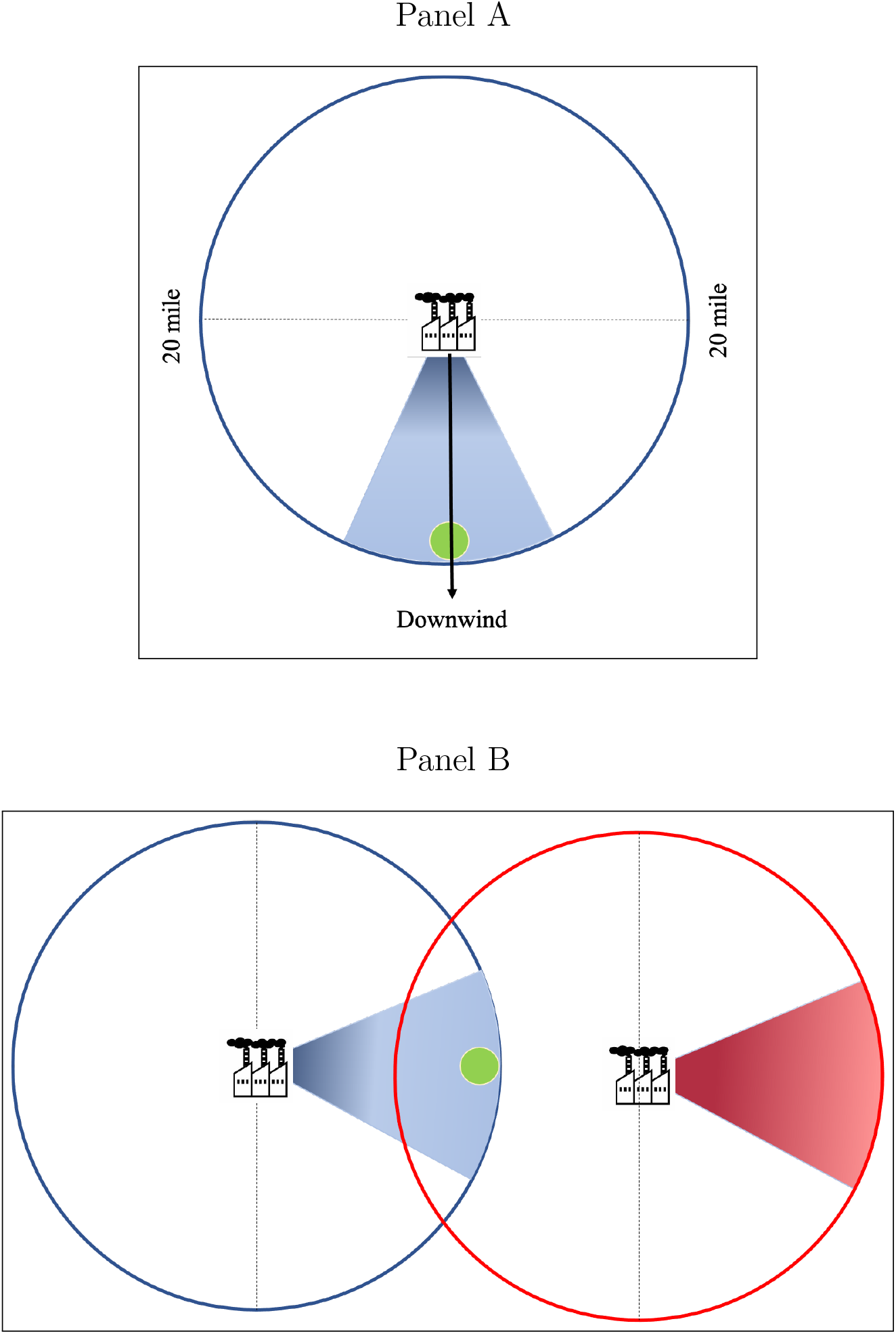
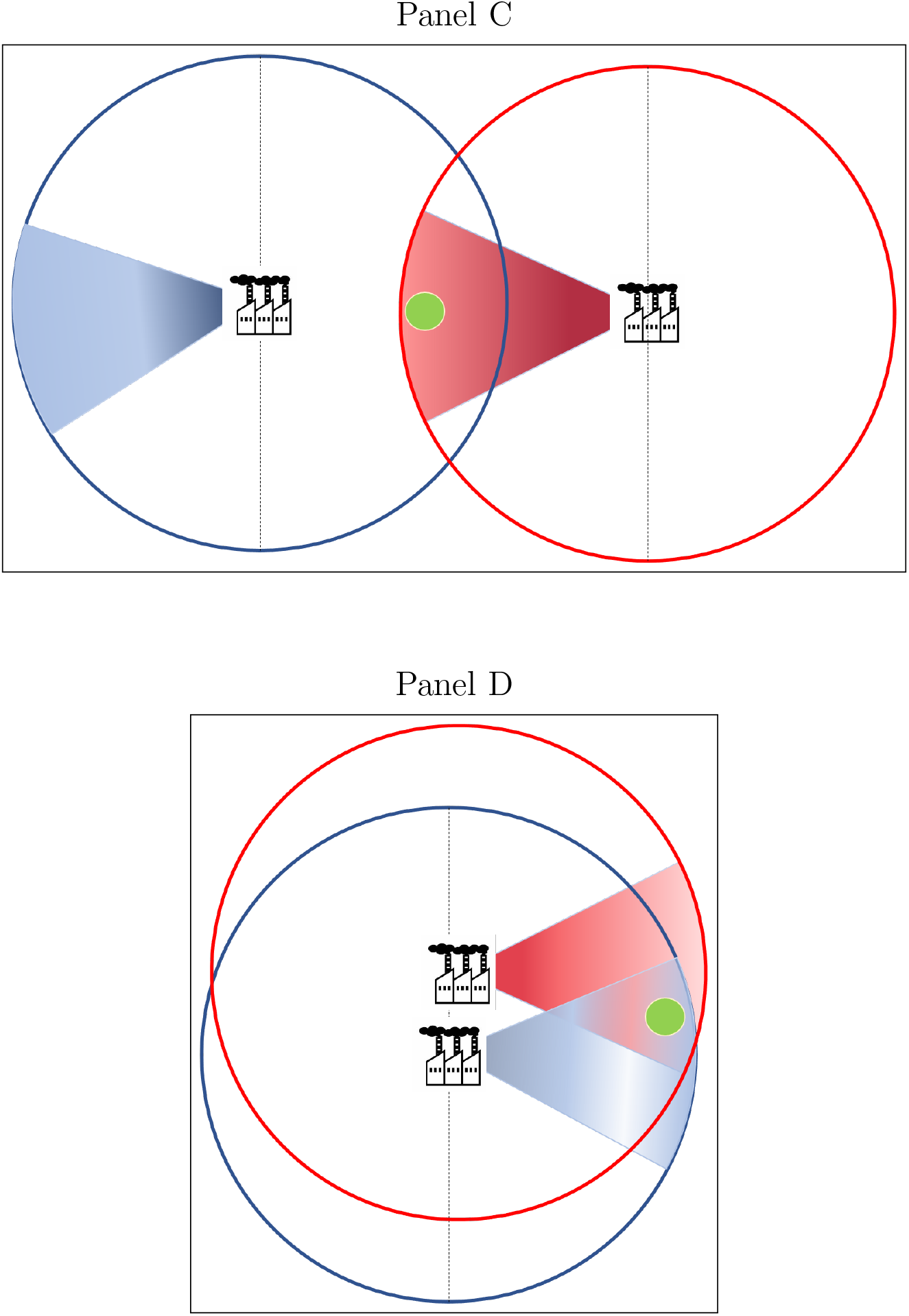
Downwind *Notes*: All panels illustrate the cases when the county whose centroid is illustrated by the green circles is considered to be downwind. In particular, the county is considered to be downwind of power plants on the day when the county’s centroid is located within 45 degrees of a ray from the power plants to wind direction, i.e., 22.5 degrees both eastward and westward. We consider all power plants within 20 miles of county centroids in constructing the dummy variable for being downwind on the daily basis. Each circle around a power plant represents the radius of 20 miles.

**Figure A.4:**
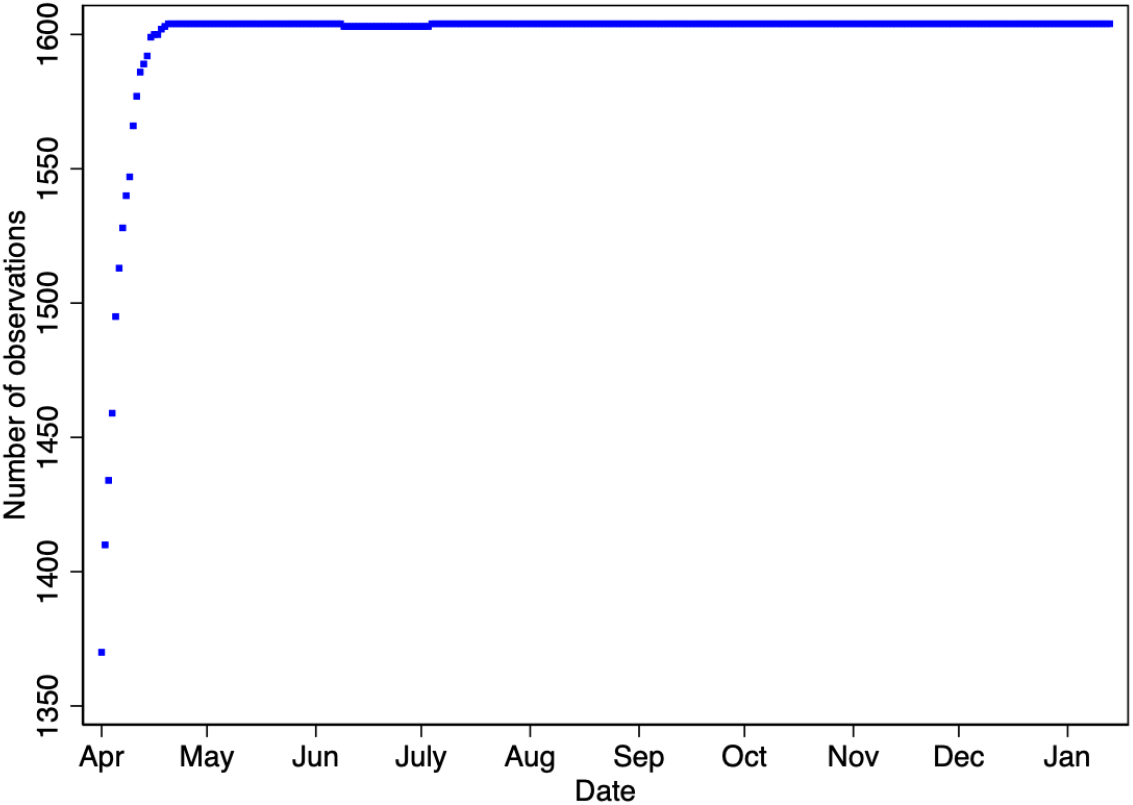
Number of Observations *Notes*: This figure plots the number of observations for each regression whose results are illustrated by Figure 3.

**Figure A.5:**
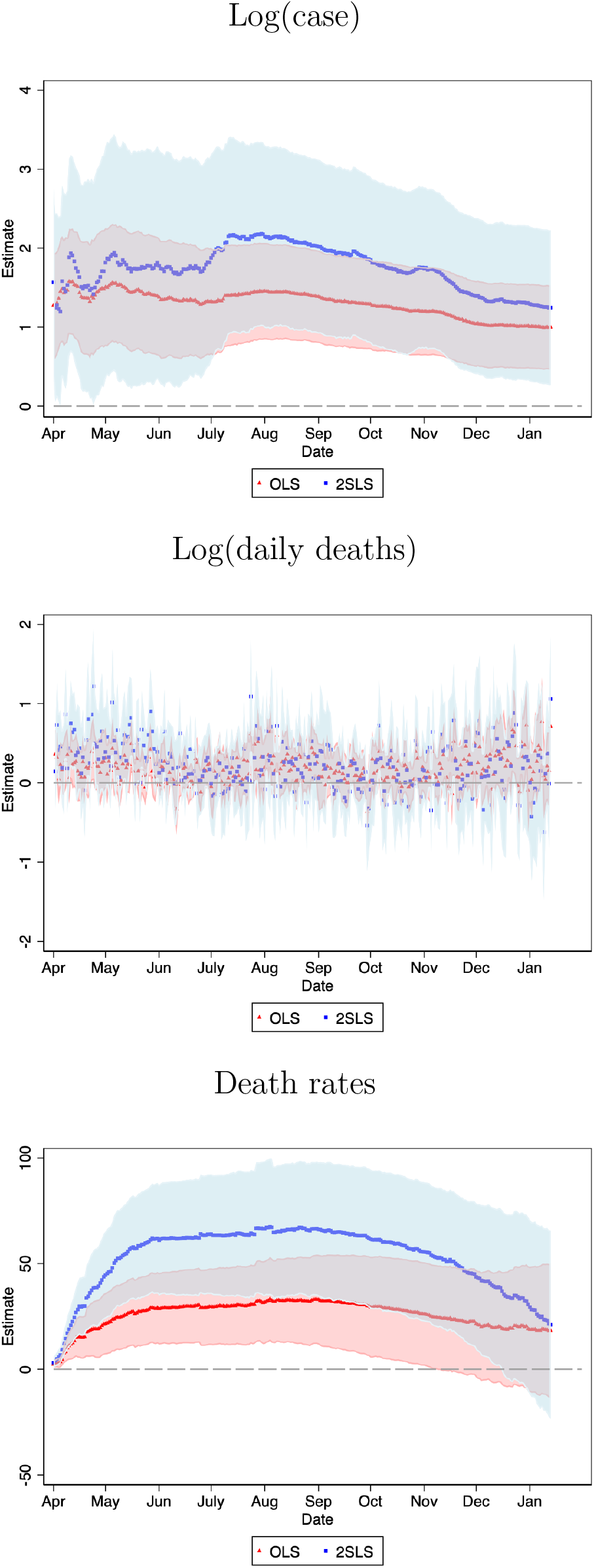
Alternative Dependent Variables *Notes*: These figures plot the coefficients and the 95% confidence intervals for each day between April 1, 2020 and January 13, 2021 estimated by OLS in red and 2SLS in blue. The dependent variables are the log of the respective number plus one in the top two panels and the mortality rate per 100,000 population in the bottom panel. The regressions control for the full set of county characteristics and state fixed effects. The number of observations is 1,604 for most regressions.

**Figure A.6:**
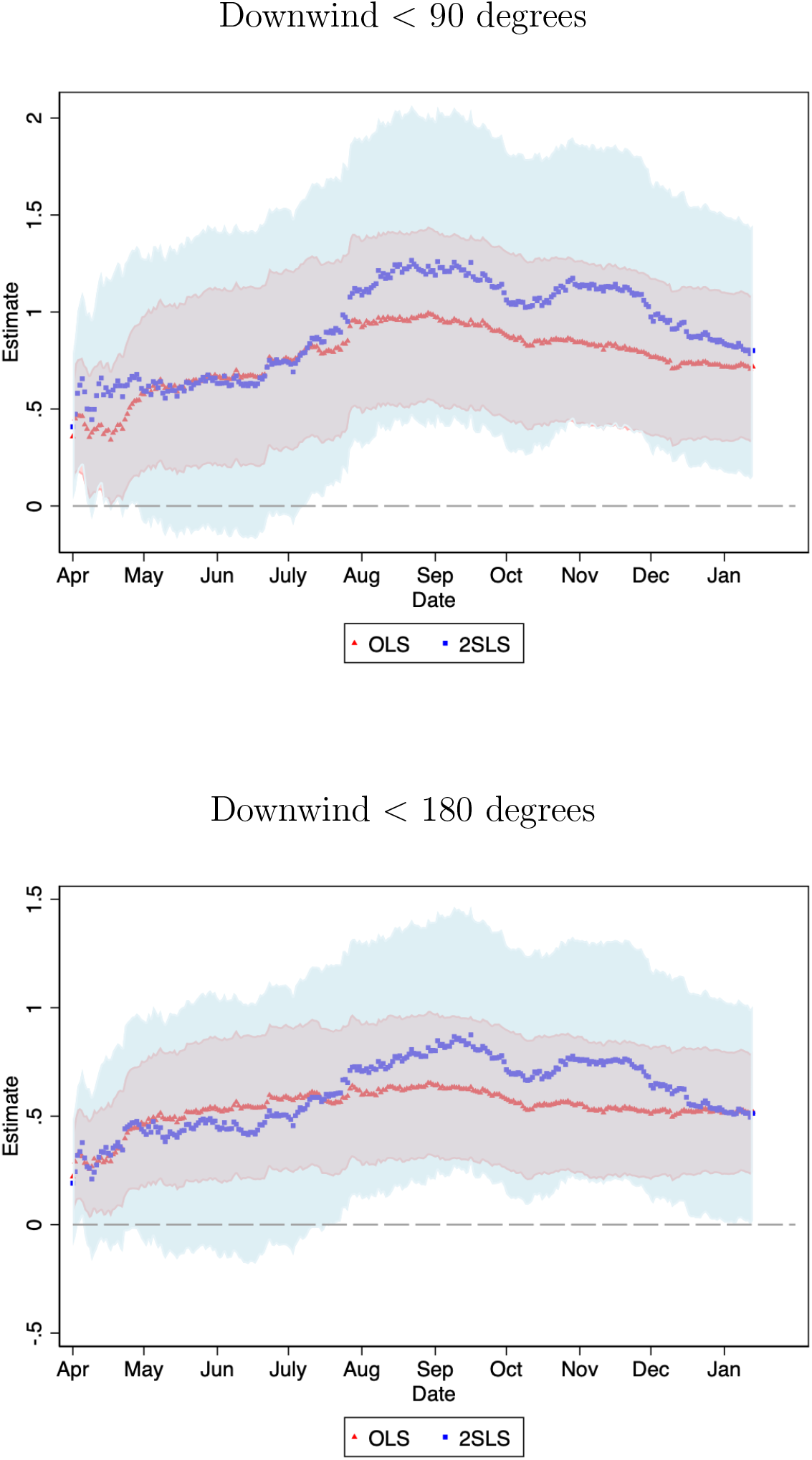
Alternative Independent Variables *Notes*: These figures plot the coefficients and the 95% confidence intervals for each day using OLS in red and 2SLS in blue for the respective independent variable between April 2020 and January 13, 2021.

**Figure A.7:**
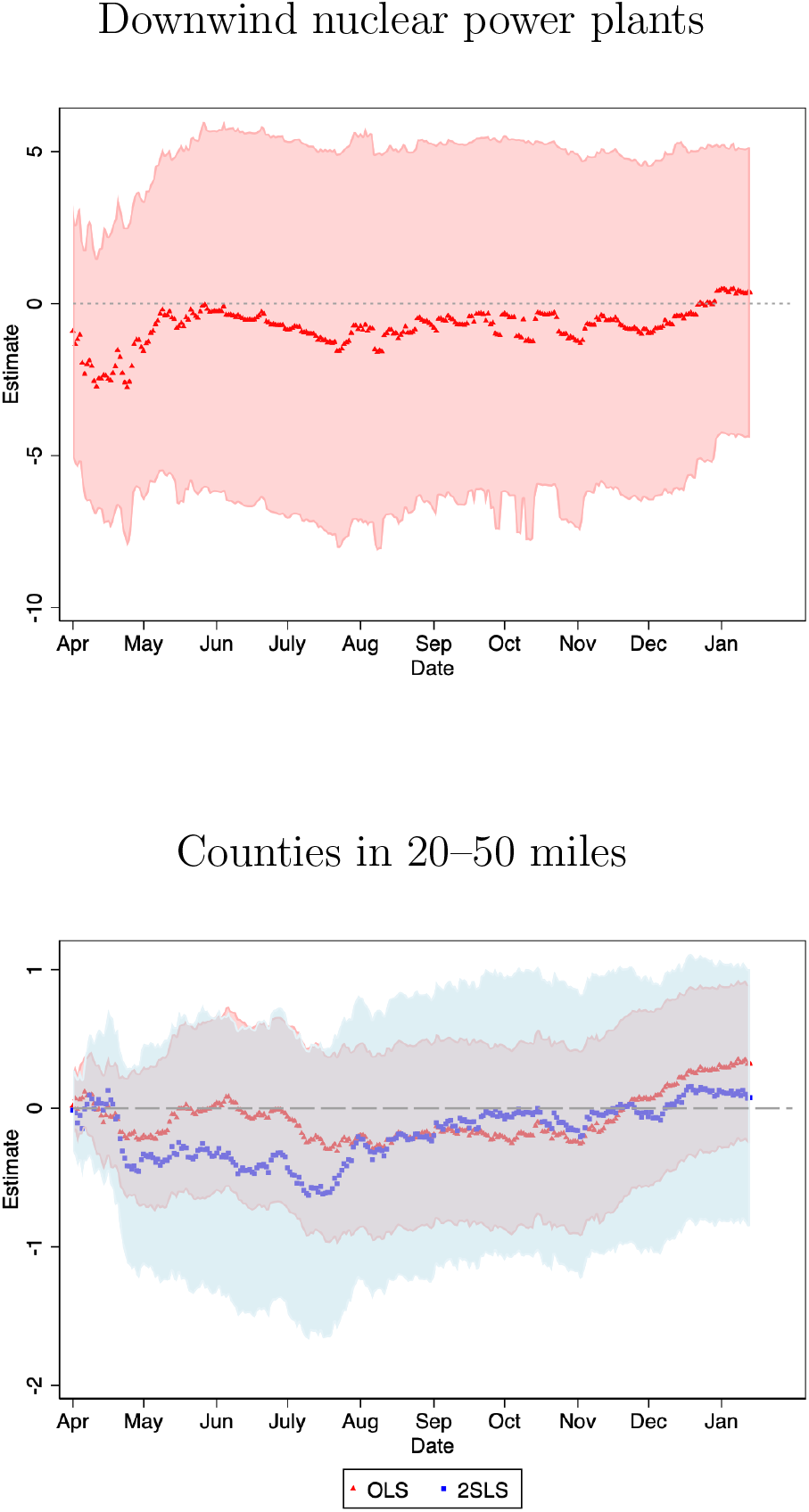
Falsification Evidence *Notes*: These figures plot the coefficients and the 95% confidence intervals for each day using OLS in red and 2SLS in blue for being downwind of nuclear power plants in the top figure and for counties in 20–50 miles away from fossil fuel power plants between April 2020 and January 13, 2021. The specification for downwind of nuclear power plants presents only OLS estimates as there are not sufficient observations to run the 2SLS analysis.

**Figure A.8:**
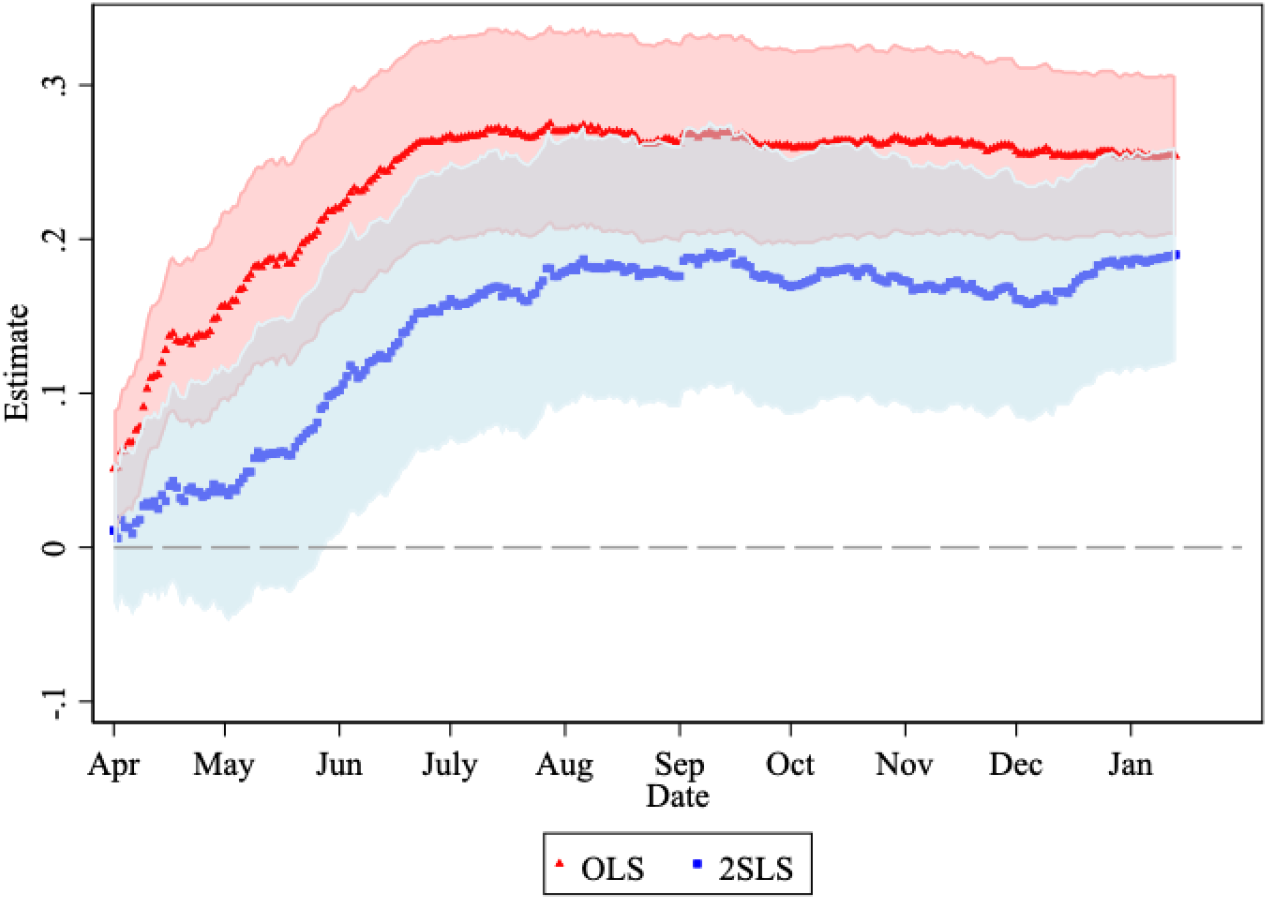
The Effects of Long-term PM2.5 Exposure on COVID-19 Mortality *Notes*: This figure plots the coefficients and the 95% confidence intervals for each day using OLS in red and 2SLS in blue with regard to the effects of long-term average PM2.5 concentrations on COVID-19 mortality for counties in 20–50 miles away from fossil fuel power plants between April 2020 and January 13, 2021.

**Table A.1:** Summary Statistics

| Variables | Mean | Std. Dev. |
| --- | --- | --- |
| *Panel A: COVID-19 as of January 13, 2021* |  |  |
| No. of deaths in sample | 190.37 | 546.72 |
| Mortality rate (per 100K) | 124.02 | 71.85 |
| No. of cases | 11,762.6 | 35,498.6 |
| *Panel B: Power plants* |  |  |
| Downwind < 45 degree | 0.135 | 0.074 |
| Downwind < 90 degree | 0.256 | 0.114 |
| Downwind < 180 degree | 0.496 | 0.150 |
| Distance to plant | 9.300 | 5.622 |
| No. of plants | 2.999 | 2.506 |
| *Panel C: County characteristics* |  |  |
| Median age | 40.235 | 4.726 |
| Gender (No. of women per 100 men) | 99.184 | 9.043 |
| Percent white | 79.626 | 16.357 |
| Percent black | 10.910 | 14.745 |
| Percent of aged 65+ | 17.139 | 4.001 |
| Population density (people/sq. miles) | 382.600 | 1,110.3 |
| Percent poverty | 14.052 | 5.567 |
| Unemployment rate | 4.049 | 1.318 |
| Median income | 57,000.0 | 15,000.0 |
| Percent without health insurance aged 65+ | 0.076 | 0.101 |
| Percent Medicare aged 65+ | 4.461 | 1.367 |
| Percent Medicaid aged 65+ | 1.135 | 0.711 |
| Percent less than a high school degree | 12.604 | 5.645 |
| Percent with a high school degree | 33.273 | 7.640 |
| Percent with a bachelor's degree + | 23.781 | 10.493 |
| Median house value | 160,000.0 | 97,000.0 |
| ICU beds | 48.357 | 135.302 |
| Daily precipitation (mm) | 3.050 | 0.869 |
| Average humidity (%) | 65.147 | 5.451 |
| Average temperature (C) | 13.451 | 4.206 |
| Average latitude | 38.126 | 4.451 |
| Average longitude | -89.099 | 11.175 |
| Total population | 170,000.0 | 440,000.0 |
| Long-term PM2.5 ($\mu\text{g}/\text{m}^3$) | 7.871 | 1.695 |
| *Panel D: Air quality monitors* |  |  |
| PM2.5 ($\mu\text{g}/\text{m}^3$) | 8.730 | 5.881 |
| Downwind < 45 degree | 0.148 | 0.355 |
| Downwind < 90 degree | 0.269 | 0.443 |
| Downwind < 180 degree | 0.511 | 0.500 |
| Distance to plant (mile) | 5.784 | 4.920 |

*Notes:* This table presents the summary statistics of key variables in the analysis. The sample includes counties in the contiguous U.S. within 20 miles of fossil fuel power plants with COVID-19 mortality data. In total, there are 1,604 counties. The levels of observations are at the county level for Panels A, B, and C, and at the daily-monitor level in Panel D.

**Table A.2:**
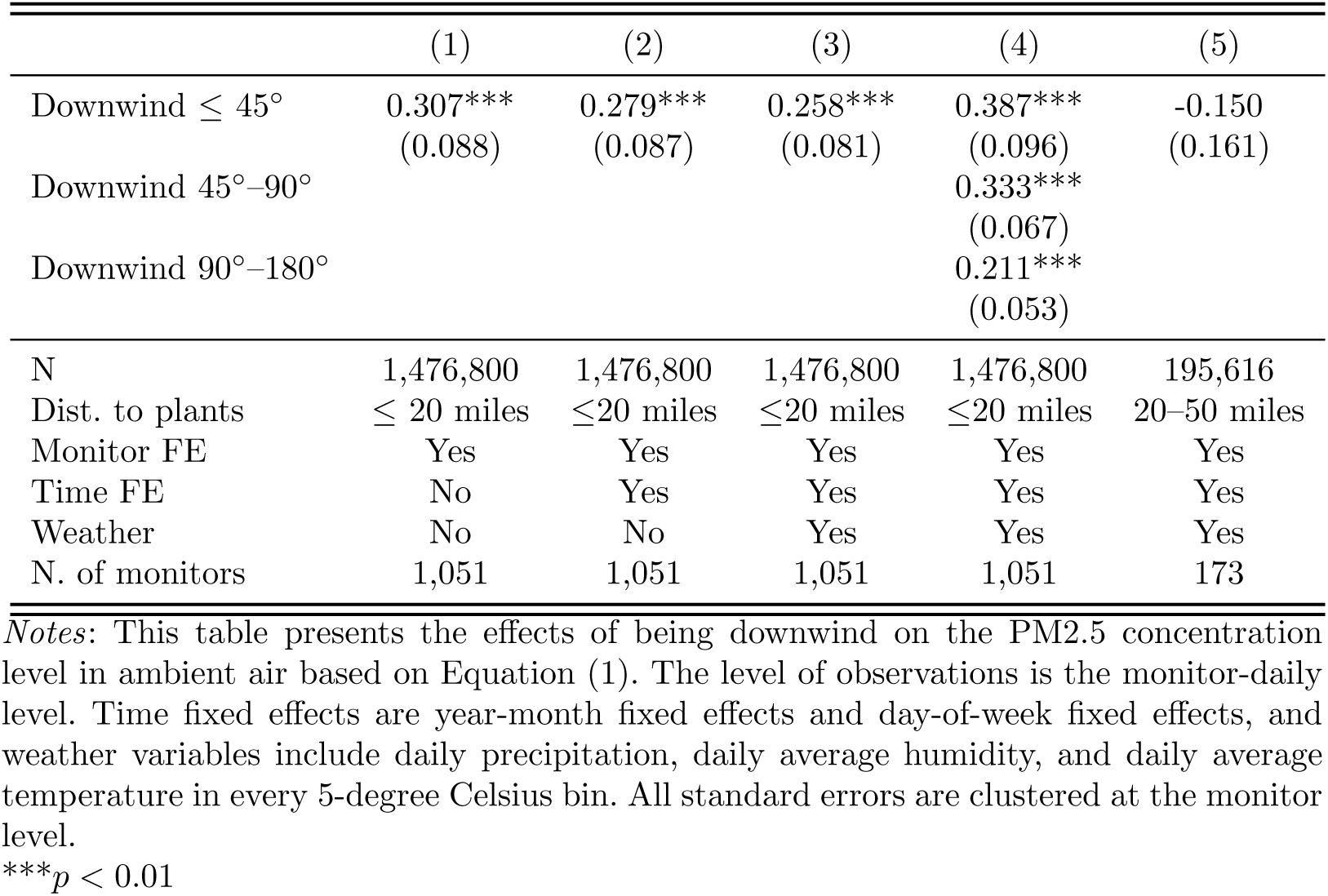
Effect of Downwind on PM2.5

|  | (1) | (2) | (3) | (4) | (5) |
| --- | --- | --- | --- | --- | --- |
| Downwind $\leq 45^\circ$ | 0.307***<br>(0.088) | 0.279***<br>(0.087) | 0.258***<br>(0.081) | 0.387***<br>(0.096) | -0.150<br>(0.161) |
| Downwind $45^\circ-90^\circ$ | | | | 0.333***<br>(0.067) | |
| Downwind $90^\circ-180^\circ$ | | | | 0.211***<br>(0.053) | |
| N | 1,476,800 | 1,476,800 | 1,476,800 | 1,476,800 | 195,616 |
| Dist. to plants | $\leq 20$ miles | $\leq 20$ miles | $\leq 20$ miles | $\leq 20$ miles | 20–50 miles |
| Monitor FE | Yes | Yes | Yes | Yes | Yes |
| Time FE | No | Yes | Yes | Yes | Yes |
| Weather | No | No | Yes | Yes | Yes |
| N. of monitors | 1,051 | 1,051 | 1,051 | 1,051 | 173 |

*Notes:* This table presents the effects of being downwind on the PM2.5 concentration level in ambient air based on Equation (1). The level of observations is the monitor-daily level. Time fixed effects are year-month fixed effects and day-of-week fixed effects, and weather variables include daily precipitation, daily average humidity, and daily average temperature in every 5-degree Celsius bin. All standard errors are clustered at the monitor level.
\*\*\* $p < 0.01$

**Table A.3:**
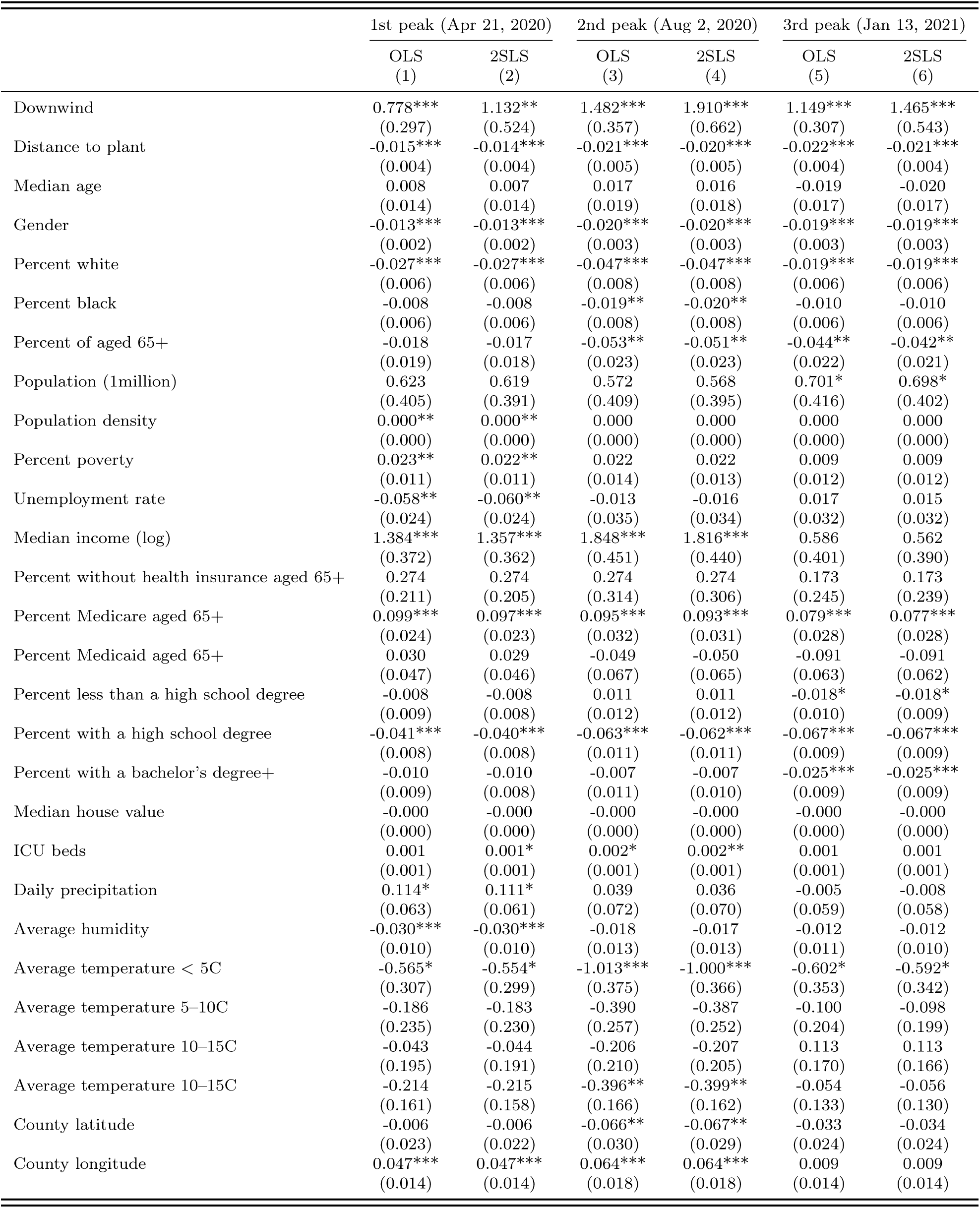
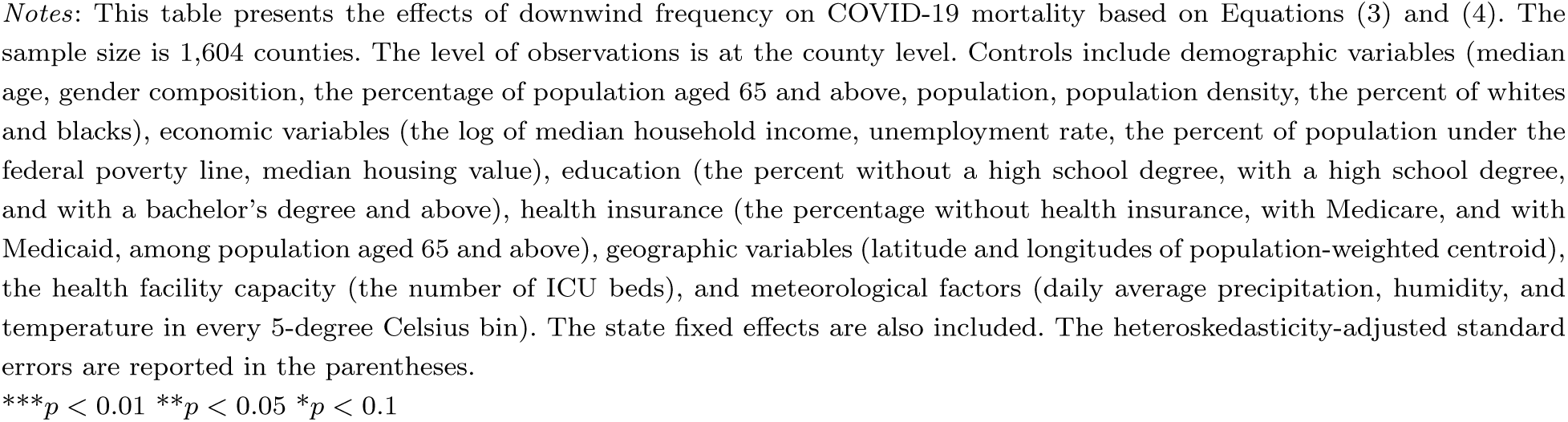
Effect of Downwind Frequency on COVID-19 Mortality

|  | 1st peak (Apr 21, 2020) |  | 2nd peak (Aug 2, 2020) |  | 3rd peak (Jan 13, 2021) |  |
| --- | --- | --- | --- | --- | --- | --- |
|  | OLS<br>(1) | 2SLS<br>(2) | OLS<br>(3) | 2SLS<br>(4) | OLS<br>(5) | 2SLS<br>(6) |
| Downwind | 0.778***<br>(0.297) | 1.132**<br>(0.524) | 1.482***<br>(0.357) | 1.910***<br>(0.662) | 1.149***<br>(0.307) | 1.465***<br>(0.543) |
| Distance to plant | -0.015***<br>(0.004) | -0.014***<br>(0.004) | -0.021***<br>(0.005) | -0.020***<br>(0.005) | -0.022***<br>(0.004) | -0.021***<br>(0.004) |
| Median age | 0.008<br>(0.014) | 0.007<br>(0.014) | 0.017<br>(0.019) | 0.016<br>(0.018) | -0.019<br>(0.017) | -0.020<br>(0.017) |
| Gender | -0.013***<br>(0.002) | -0.013***<br>(0.002) | -0.020***<br>(0.003) | -0.020***<br>(0.003) | -0.019***<br>(0.003) | -0.019***<br>(0.003) |
| Percent white | -0.027***<br>(0.006) | -0.027***<br>(0.006) | -0.047***<br>(0.008) | -0.047***<br>(0.008) | -0.019***<br>(0.006) | -0.019***<br>(0.006) |
| Percent black | -0.008<br>(0.006) | -0.008<br>(0.006) | -0.019**<br>(0.008) | -0.020**<br>(0.008) | -0.010<br>(0.006) | -0.010<br>(0.006) |
| Percent of aged 65+ | -0.018<br>(0.019) | -0.017<br>(0.018) | -0.053**<br>(0.023) | -0.051**<br>(0.023) | -0.044**<br>(0.022) | -0.042**<br>(0.021) |
| Population (1million) | 0.623<br>(0.405) | 0.619<br>(0.391) | 0.572<br>(0.409) | 0.568<br>(0.395) | 0.701*<br>(0.416) | 0.698*<br>(0.402) |
| Population density | 0.000**<br>(0.000) | 0.000**<br>(0.000) | 0.000<br>(0.000) | 0.000<br>(0.000) | 0.000<br>(0.000) | 0.000<br>(0.000) |
| Percent poverty | 0.023**<br>(0.011) | 0.022**<br>(0.011) | 0.022<br>(0.014) | 0.022<br>(0.013) | 0.009<br>(0.012) | 0.009<br>(0.012) |
| Unemployment rate | -0.058**<br>(0.024) | -0.060**<br>(0.024) | -0.013<br>(0.035) | -0.016<br>(0.034) | 0.017<br>(0.032) | 0.015<br>(0.032) |
| Median income (log) | 1.384***<br>(0.372) | 1.357***<br>(0.362) | 1.848***<br>(0.451) | 1.816***<br>(0.440) | 0.586<br>(0.401) | 0.562<br>(0.390) |
| Percent without health insurance aged 65+ | 0.274<br>(0.211) | 0.274<br>(0.205) | 0.274<br>(0.314) | 0.274<br>(0.306) | 0.173<br>(0.245) | 0.173<br>(0.239) |
| Percent Medicare aged 65+ | 0.099***<br>(0.024) | 0.097***<br>(0.023) | 0.095***<br>(0.032) | 0.093***<br>(0.031) | 0.079***<br>(0.028) | 0.077***<br>(0.028) |
| Percent Medicaid aged 65+ | 0.030<br>(0.047) | 0.029<br>(0.046) | -0.049<br>(0.067) | -0.050<br>(0.065) | -0.091<br>(0.063) | -0.091<br>(0.062) |
| Percent less than a high school degree | -0.008<br>(0.009) | -0.008<br>(0.008) | 0.011<br>(0.012) | 0.011<br>(0.012) | -0.018*<br>(0.010) | -0.018*<br>(0.009) |
| Percent with a high school degree | -0.041***<br>(0.008) | -0.040***<br>(0.008) | -0.063***<br>(0.011) | -0.062***<br>(0.011) | -0.067***<br>(0.009) | -0.067***<br>(0.009) |
| Percent with a bachelor's degree+ | -0.010<br>(0.009) | -0.010<br>(0.008) | -0.007<br>(0.011) | -0.007<br>(0.010) | -0.025***<br>(0.009) | -0.025***<br>(0.009) |
| Median house value | -0.000<br>(0.000) | -0.000<br>(0.000) | -0.000<br>(0.000) | -0.000<br>(0.000) | -0.000<br>(0.000) | -0.000<br>(0.000) |
| ICU beds | 0.001<br>(0.001) | 0.001*<br>(0.001) | 0.002*<br>(0.001) | 0.002**<br>(0.001) | 0.001<br>(0.001) | 0.001<br>(0.001) |
| Daily precipitation | 0.114*<br>(0.063) | 0.111*<br>(0.061) | 0.039<br>(0.072) | 0.036<br>(0.070) | -0.005<br>(0.059) | -0.008<br>(0.058) |
| Average humidity | -0.030***<br>(0.010) | -0.030***<br>(0.010) | -0.018<br>(0.013) | -0.017<br>(0.013) | -0.012<br>(0.011) | -0.012<br>(0.010) |
| Average temperature < 5C | -0.565*<br>(0.307) | -0.554*<br>(0.299) | -1.013***<br>(0.375) | -1.000***<br>(0.366) | -0.602*<br>(0.353) | -0.592*<br>(0.342) |
| Average temperature 5–10C | -0.186<br>(0.235) | -0.183<br>(0.230) | -0.390<br>(0.257) | -0.387<br>(0.252) | -0.100<br>(0.204) | -0.098<br>(0.199) |
| Average temperature 10–15C | -0.043<br>(0.195) | -0.044<br>(0.191) | -0.206<br>(0.210) | -0.207<br>(0.205) | 0.113<br>(0.170) | 0.113<br>(0.166) |
| Average temperature 10–15C | -0.214<br>(0.161) | -0.215<br>(0.158) | -0.396**<br>(0.166) | -0.399**<br>(0.162) | -0.054<br>(0.133) | -0.056<br>(0.130) |
| County latitude | -0.006<br>(0.023) | -0.006<br>(0.022) | -0.066**<br>(0.030) | -0.067**<br>(0.029) | -0.033<br>(0.024) | -0.034<br>(0.024) |
| County longitude | 0.047***<br>(0.014) | 0.047***<br>(0.014) | 0.064***<br>(0.018) | 0.064***<br>(0.018) | 0.009<br>(0.014) | 0.009<br>(0.014) |

*Notes:* This table presents the effects of downwind frequency on COVID-19 mortality based on Equations (3) and (4). The sample size is 1,604 counties. The level of observations is at the county level. Controls include demographic variables (median age, gender composition, the percentage of population aged 65 and above, population, population density, the percent of whites and blacks), economic variables (the log of median household income, unemployment rate, the percent of population under the federal poverty line, median housing value), education (the percent without a high school degree, with a high school degree, and with a bachelor's degree and above), health insurance (the percentage without health insurance, with Medicare, and with Medicaid, among population aged 65 and above), geographic variables (latitude and longitudes of population-weighted centroid), the health facility capacity (the number of ICU beds), and meteorological factors (daily average precipitation, humidity, and temperature in every 5-degree Celsius bin). The state fixed effects are also included. The heteroskedasticity-adjusted standard errors are reported in the parentheses.
\*\*\* $p < 0.01$ \*\* $p < 0.05$ \* $p < 0.1$

**Table A.4:** Robustness checks and falsification tests

| Specification | 1st peak (Apr 21, 2020) |  | 2nd peak (Aug 2, 2020) |  | 3rd peak (Jan 13, 2021) |  |
| --- | --- | --- | --- | --- | --- | --- |
|  | OLS<br>(1) | 2SLS<br>(2) | OLS<br>(3) | 2SLS<br>(4) | OLS<br>(5) | 2SLS<br>(6) |
| *Panel A: Robustness* |  |  |  |  |  |  |
| Log(case) as outcome | 1.366***<br>(0.366) | 1.528**<br>(0.688) | 1.454***<br>(0.311) | 2.136***<br>(0.588) | 0.998***<br>(0.272) | 1.248**<br>(0.499) |
| Log(daily deaths) as outcome | 0.179<br>(0.194) | 0.803**<br>(0.367) | 0.673***<br>(0.198) | 0.653**<br>(0.261) | 0.711***<br>(0.238) | 1.057***<br>(0.396) |
| Death rate as outcome | 17.899***<br>(6.122) | 34.540***<br>(8.109) | 32.235***<br>(10.019) | 67.141***<br>(16.318) | 18.240<br>(16.264) | 21.037<br>(22.661) |
| Consider 90 degrees downwind | 0.418**<br>(0.190) | 0.620*<br>(0.319) | 0.930***<br>(0.229) | 1.105***<br>(0.404) | 0.718***<br>(0.192) | 0.801**<br>(0.330) |
| Consider 180 degrees downwind | 0.353**<br>(0.137) | 0.359<br>(0.239) | 0.600***<br>(0.167) | 0.710**<br>(0.301) | 0.520***<br>(0.142) | 0.513**<br>(0.252) |
| Control for short-term downwind frequency | 0.927***<br>(0.359) | 1.446**<br>(0.630) | 1.565***<br>(0.443) | 2.541***<br>(0.814) | 1.101***<br>(0.315) | 1.483***<br>(0.550) |
| Control for number of power plants | 0.694**<br>(0.298) | 0.973*<br>(0.533) | 1.367***<br>(0.358) | 1.692**<br>(0.674) | 1.089***<br>(0.308) | 1.341**<br>(0.553) |
| Control for number of TRI firms | 0.755**<br>(0.295) | 1.144**<br>(0.523) | 1.429***<br>(0.351) | 1.900***<br>(0.654) | 1.106***<br>(0.303) | 1.448***<br>(0.539) |
| *Panel B: Falsification* |  |  |  |  |  |  |
| Downwind nuclear | -1.769<br>(2.495) |  | -0.877<br>(3.199) |  | 0.363<br>(2.396) |  |
| Counties in 20–50 miles | -0.174<br>(0.203) | -0.218<br>(0.327) | -0.192<br>(0.340) | -0.224<br>(0.519) | 0.321<br>(0.292) | 0.077<br>(0.472) |

*Notes:* This table presents evidence from the robustness checks of the main analysis against various alternative specifications in Panel A and evidence from falsification tests in Panel B. All regressions include the state fixed effects and all controls from the main analysis. The specification for downwind of nuclear power plants presents only OLS estimates because there are not sufficient observations to run the 2SLS analysis. The heteroskedasticity-adjusted standard errors are reported in the parentheses. \*\*\* $p < 0.01$ \*\* $p < 0.05$

**Table A.5:** Heterogeneities in the effects of downwind frequency on COVID-19 mortality

|  | Relative to median |  |  |  |  |  |
| --- | --- | --- | --- | --- | --- | --- |
|  | Below | Above | Below | Above | Below | Above |
|  | 1st peak (Apr 21, 2020)<br>(1) | (2) | 2nd peak (Aug 2, 2020)<br>(3) | (4) | 3rd peak (Jan 13, 2021)<br>(5) | (6) |
| Gender (No. of women per 100 men) | 0.844*<br>(0.463) | 0.307<br>(0.373) | 1.752***<br>(0.502) | 1.036**<br>(0.513) | 1.356***<br>(0.416) | 0.831**<br>(0.421) |
| Percent of black | 0.655*<br>(0.338) | 0.266<br>(0.479) | 1.092**<br>(0.443) | 1.020*<br>(0.556) | 0.821**<br>(0.370) | 0.719*<br>(0.435) |
| Percent of old | 0.528<br>(0.408) | 0.731**<br>(0.365) | 1.080**<br>(0.499) | 1.460***<br>(0.519) | 0.904**<br>(0.424) | 0.841**<br>(0.392) |
| Poverty rate | 0.544<br>(0.438) | 0.698*<br>(0.374) | 0.746<br>(0.504) | 1.660***<br>(0.491) | 0.859**<br>(0.426) | 1.048**<br>(0.422) |
| Percent of no health insurance among 65+ | -0.200<br>(0.349) | 1.128**<br>(0.442) | 0.473<br>(0.494) | 1.641***<br>(0.489) | 0.413<br>(0.388) | 1.235***<br>(0.420) |
| Percent with less than a high school degree | 0.431<br>(0.394) | 0.943**<br>(0.388) | 0.677<br>(0.490) | 1.969***<br>(0.489) | 0.243<br>(0.407) | 1.595***<br>(0.406) |

*Notes:* This table reports the estimated effects of downwind frequency on COVID-19 mortality for various subsample of counties determined by being above or below the median of the respective county characteristics specified in each row. The analysis is based on OLS because there are not sufficient observations to run 2SLS. The analysis includes the set of county characteristics and state fixed effects. The heteroskedasticity-adjusted standard errors are reported in the parentheses.
\*\*\* $p < 0.01$ , \*\* $p < 0.05$ , \* $p < 0.1$

**Table A.6:**
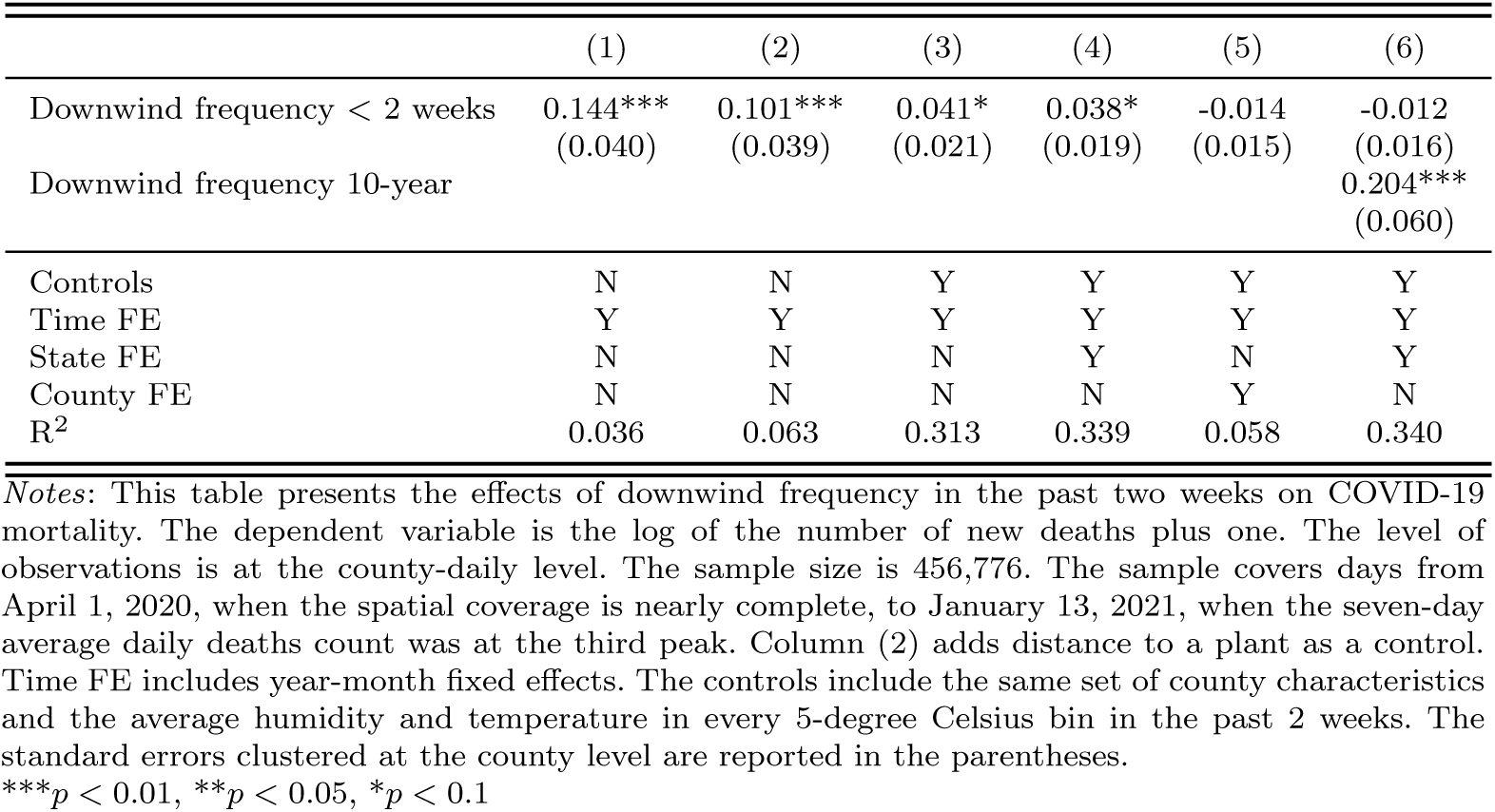
Effect of recent downwind frequency on COVID-19 mortality

## Footnotes

1 See an extensive literature reviewed in Online Appendix A.

2 Note that van Donkelaar et al. (2019) construct such data for 2000–2016. To be consistent with our study period, we focus on the period of 2010–2016, yet the long-term average PM2.5 concentrations at the county level in 2000–2016 and in 2010–2016 are highly correlated with the correlation coefficient of 0.98. Thus, the average PM2.5 concentrations in 2017–2019, if they were available, would likely be similar.

3 See Online Appendix Section B for detailed description.

4 We also drop New York City (NYC) due to an obvious reporting error in the number; specifically, the cumulative number of deaths falls from 23,689 to 3,170 on August 31, 2020, and starts increasing from there onward, reaching cumulative counts of only 3,586 even on January 31, 2021.

5 The data on the population-weighted county centroids are obtained from U.S. Census Bureau (2020).

6 https://www.epa.gov/pm-pollution/particulate-matter-pm-basics

7 When there are multiple plants within 20 miles of an air quality monitor, the monitor is considered to be downwind when it is downwind of at least one power plant.

8 While taking the logarithm is appropriate to address both outliers in the number of deaths and the exponential increase in COVID deaths (Bursztyn et al. 2020), we also consider mortality rates as an alternative outcome.

9 See Online Appendix A.3 for illustrative figures.

10 Additional concerns that are addressed by the 2SLS analysis are that i) there is a measurement error in the exposure measure and ii) counties that accommodate power plants are relatively more likely to be downwind than those without, although we find no association between downwind frequency and the number of fossil fuel power plants within the county, enhancing exogeneity of the wind direction with respect to the county centroid.

11 Deryugina et al. (2019) uses the daily wind direction in the county as an instrument for PM2.5 whose relationships are allowed to vary across geography in estimating the effect of air pollution on adult mortality.

12 Online Appendix Figure A.2 illustrates the distributions of these groups. Note that there is little theoretical guidance as to how many clusters are optimal except than that the monotonicity assumption must hold within each cluster. We chose 90 as these clusters classify the entire country into spatially proximate groups, within which the effects of bearing angle on downwind frequency are similar as evidenced in the strong first stage results.

13 See Online Appendix C for further discussions.

14 Note that the number of monitors is substantially lower in these areas although they cover more than 5 times the land area. Since the EPA typically places monitors near pollution sources, e.g. power plants, the number of monitors by itself reflects recognition that the population within 20 miles of power plants is at substantially greater risk.

15 See Online Appendix Figure A.1 for the trends in COVID-19 in the U.S.

16 See Online Appendix Table A.3 for all other coefficients and Section D.1 for further discussions.

17 An alternative way to interpret these estimates is that each additional 10 days spent downwind in the last 10 years led to 0.10, 1.17, or 1.73 more deaths at the first, second, and third peak, respectively.

18 We additionally examined whether the effects differ between coal-fired power plants and natural gas-fired power plants, and we consistently find larger effects for natural gas. This may sound counterintuitive given that coal-fired power plants emit a larger amount of emissions. Our results possibly reflect that 68% of the county centroids in our sample are situated closer to natural gas-fired power plants than coal-fired power plants. Natural gas-fired power plants tend to be located near county centroids than coal-fired power plants possibly due to the concerns of larger emissions from coal-fired power plants. It is also important to note that we need to focus only on the nearest power plant for this analysis, while many county centroids are in the neighborhood of multiple power plants with the average of three.

19 See Online Appendix Table A.4 and Section D.2 for further discussions.

20 See Online Appendix Table A.6 and Section D.5 for further discussions

21 See Online Appendix Table A.4 and Section D.3 for further discussions.

22 See Online Appendix Table A.5 and Section D.4 for further discussions.

23 On a separate note, we find that long-term average PM2.5 concentrations are significantly associated with downwind frequency at the county level. Based on the similar model with the extensive set of county characteristics and state fixed effects, the coefficients (standard errors) of OLS and IV are 1.291 (0.319) and 1.423 (0.589), respectively.

24 https://www.un.org/press/en/2020/sgsm20063.doc.htm

1 In the U.S,. the Clean Air Act required the Environmental Protection Agency (EPA) to set health-based National Ambient Air Quality Standards for pollutants typically emitted from fossil fuel power plants. The Clean Air Act Amendments of 1990 established the Acid Rain Program, the allowance market system of SO_2_ and NO_X_ from coal-fired power plants. In 2011, the EPA introduced the Mercury and Air Toxics Standards, the first set of federal mandates to limit mercury and other hazardous pollutants from coal- and oil-fired power plants.

2 Note that these sample means are not simply inferred from the angle fraction of a circle because the sample excludes even contiguous counties of plants when their centroids are located beyond 20 miles of plants.

3 Inevitably, we need to limit this analysis to the pair of monitors and the nearest power plants, and thereby having a disadvantage over the main analysis of confounding effects from other power plants in the neighborhood.

4 Note that when the mortality rates are used as the outcome, we follow the convention that all regressions are weighted by the county population, as counties with greater population allow precise estimates of averages.

5 The corresponding OLS estimates suggest that counties with average downwind frequency experienced 150.0 (39.4%), 1053.8 (44.3%), 2719.9 (23.1%) more cases, 0.028 (2.65%), 0.031 (13.0%), 0.319 (14.0%) more daily new deaths, and 2.42 (46.0%), 4.35 (14.9%), 2.46 (1.99%) more deaths per 100,000 population at the first, second, and third peak days respectively than completely upwind counties within the same distance from plants.

6 The analysis controls for downwind frequency of fossil fuel power plants in order to account for spatial correlations between nuclear power plants and fossil fuel power plants.

7 The specification for downwind of nuclear power plants presents only OLS estimates as there are not sufficient observations to run the 2SLS analysis.

8 The analysis is based on OLS because there are not sufficient observations to run 2SLS.

